# Cognitive bias modification for emotional facial expressions modifies neural mechanisms in individuals taking antidepressant medication: a Randomised Controlled Trial

**DOI:** 10.64898/2026.01.27.26344720

**Authors:** Charlotte M Crisp, Sean James Fallon, Alison Burns, Rumeysa Kuruoğlu, Jennifer Ferrar, Nicola Wiles, David Kessler, Marcus R Munafò, Ian S Penton-Voak

**Affiliations:** Population Health Sciences, Bristol Medical School, University of Bristol, Bristol, UK; NIHR Bristol Biomedical Research Centre, UK; School of Psychology, University of Plymouth, UK; Bristol Trials Centre, Population Health Sciences, University of Bristol, UK; School of Psychological Science, University of Bristol, UK; Centre for Academic Mental Health, Population Health Sciences, University of Bristol, UK; Vice Chancellor’s Office, University of Bath, UK

## Abstract

**Background:** Antidepressants exert their therapeutic effects through ameliorating negative emotional biases that underpin depression. However, therapeutic gains may depend upon restructuring how emotional information is processed. This can be achieved through Cognitive Bias Modification (CBM), a technique for positively shifting recognition of emotional facial expressions. Here, we examined how CBM modifies emotional processing circuits in individuals with depression who were taking Selective Serotonin Reuptake Inhibitors (SSRIs).

**Methods:** A double-blind Randomised Controlled Trial was conducted in 84 participants with depression who had recently started SSRI medication. Participants received five sessions of ‘active’ or ‘sham’ CBM over one week before fMRI scanning where they viewed emotional faces (happy, fearful, sad).

**Results:** Across all emotional expressions, greater Blood Oxygen Level Dependent (BOLD) signal was observed in the inferior occipital gyrus for the active compared to sham CBM. Emotional-specific effects were observed in the medial Prefrontal Cortex (mPFC), with reduced BOLD signal in the active (compared to sham) group for viewing happy vs. fearful faces. Changes in BOLD signal were also associated with individual differences in response to CBM. Enhanced functional connectivity between mPFC and right Dorsolateral Prefrontal Cortex (rDLPFC) predicted improvement in depressive symptoms four weeks later.

**Conclusions:** These results indicate that CBM modifies the neural circuits involved in emotion processing in people with depression currently taking antidepressants. Converting these changes in emotional perception to improved depressive symptoms was related to changing mPFC-rDLPFC connectivity. Future trials are needed to test CBM’s clinical utility as a simple, affordable and accessible adjunct therapy for depression.

## Introduction

Major Depressive Disorder (MDD) affects 5–17% of people during their lifetime and is the leading global cause of disability (Bains & Abdijadid, 2025). Up to 30% of individuals do not respond to antidepressants, and relapse rates are high (Al-Harbi, 2012). Combining antidepressant medication with psychotherapies like Cognitive Behavioural Therapy (CBT) improves outcomes, but access is limited due to high cost and long waiting lists. Novel treatment approaches for depression are therefore needed.

Negative emotional biases - preferential attention to, interpretation of, and recall of negative over positive emotional information - are implicated in the onset and maintenance of depression. Neurocognitive models suggest these biases may contribute causally to depression and are therefore a treatment target (Ian S. Penton-Voak, Munafò, & Looi, 2017) as they are observed in at-risk populations (Chan, Goodwin, & Harmer, 2007; Horne, Marr- Phillips, Jawaid, Gibson, & Norbury, 2017), normalised by antidepressants (Warren, Pringle, & Harmer, 2015; Zhang et al., 2020), and predict relapse (Ruhe et al., 2019). Treatments that modify biases in processing emotional faces (e.g., CBT, Selective Serotonin Reuptake Inhibitors; SSRIs) improve symptoms (Roiser, Elliott, & Sahakian, 2012), with changes to emotional biases preceding mood improvements and predicting recovery (Warren et al., 2015). This suggests a ‘virtuous cycle’ where more positive interpretations of emotions may improve mood through better social interactions over time (Harmer, Goodwin, & Cowen, 2009).

Functional MRI (fMRI) studies have associated negative emotional biases with altered activity in subcortical regions such as amygdala, ventral striatum and insula (Fu et al., 2004). In depression, studies have reported amygdala hyperactivity (increased BOLD signal) to negative emotions (e.g., fearful faces) and reduced response to positive emotions (e.g., happy faces) alongside attenuated top-down control from prefrontal regions (medial Prefrontal Cortex (mPFC), dorsal Anterior Cingulate Cortex (dACC) and Dorsolateral Prefrontal Cortex (DLPFC) (Disner, Beevers, Haigh, & Beck, 2011; Victor, Furey, Fromm, Ohman, & Drevets, 2010). Visual processing areas (e.g., occipital cortex, fusiform gyrus) also show altered activity in response to emotional faces (Li et al., 2013). Importantly, there is evidence that good antidepressant treatment response is associated with a reduction in BOLD signal (in amygdala, thalamus, insula and ACC) to negative emotions, suggesting bias correction may be a mechanism of antidepressant drug action (Ma, 2015; McMakin et al., 2012).

Cognitive Bias Modification (CBM) is a scalable, low-cost online behavioural intervention to improve emotional biases. Our previous work developed a CBM technique that shifts the threshold for categorising ambiguous emotional faces toward ‘happy’ using personalised feedback. CBM robustly improves emotion categorisation in clinical and non-clinical populations (R. Kuruoğlu, Attwood, & Penton-Voak, 2025; Suddell et al., 2021; Van Meter, Stoddard, Penton-Voak, & Munafò, 2021), with effects sustained after four weeks and generalising to other face stimuli (Dalili, Schofield-Toloza, Munafò, & Penton-Voak, 2017). Whilst the effects of CBM on depressive symptoms are less robust, CBM may enhance anhedonia measures, positive affect, quality of life, and perceived treatment effectiveness (I. S. Penton-Voak, Bate, Lewis, & Munafò, 2012; Peters et al., 2017; Suddell et al., 2021). In our previous fMRI study of participants reporting depressive symptoms but not using pharmacotherapy (non-clinical sample), active CBM increased BOLD signal in left amygdala and mPFC in response to happy faces, compared to sham CBM. This suggests CBM enhances perception of positive emotions, complementing SSRIs which affect responses to both negative and positive emotions (Ma, 2015; Warren et al., 2015). Combining CBM with SSRIs may therefore be particularly effective in improving emotional biases, but mechanistic studies of this are rare.

This study investigated the neural mechanisms underpinning CBM as an adjunct therapy to SSRIs in individuals with depression recruited from primary care who had recently started SSRIs. Based on our prior findings (I. S. Penton-Voak et al., 2021), we hypothesised that active CBM would positively shift emotion categorisation and increase response to positive emotions (happy-sad and happy-fear contrasts) in bilateral amygdala and mPFC, compared to sham CBM. We also explored changes in a wider network involved in attending to and regulating responses to emotional faces (e.g., occipital cortex, dACC, DLPFC) and associations with changes in depressive symptoms.

## Materials and Methods

### Study Design

A two-parallel group randomised controlled trial where participants, researchers and analysts were blind to treatment allocation. The study was based at the University of Bristol (UK). Participants were unblinded at the end of the study.

### Ethics

Ethics approval was obtained from the NHS Health and Care Research Wales (REC reference number 20/LO/1118). Health Research Authority (HRA) approval was also obtained. The trial was registered on the ISRCTN: https://www.isrctn.com/ISRCTN37448835 (12/11/2020) and the data analysis plan was pre-registered on the Open Science Framework (OSF): https://doi.org/10.17605/OSF.IO/FYUJN.

### Participants

Participants were adults (aged 18-55) with a new or first episode of depression defined as starting a course of antidepressants within the six months prior to randomisation. Participants were only recruited if they were on an SSRI medication (including sertraline, citalopram, dapoxetine, escitalopram, fluoxetine, fluvoxamine, paroxetine, zimelidine or vortioxetine) and had low mood (scored > 10 on Patient Health Questionnaire; PHQ-9 (Kroenke, Spitzer, & Williams, 2001)). Participants were excluded if they had other diagnosed psychiatric conditions (e.g., bipolar disorder, schizophrenia, alcohol/substance dependency), were receiving other high intensity psychological therapies for depression or anxiety (within the last six months) or had any contraindications for MRI scanning (see full list of inclusion and exclusion in Supplementary Materials). Eligible participants were asked not to take recreational drugs, or drink alcohol, for 72 hours and 24 hours prior to each test session, respectively.

### Recruitment

Eighty-four participants were recruited over a period of 29 months (October 2021 to February 2024). There were four recruitment streams: 1) In-consultation with General Practitioners (GPs) during primary care appointments; 2) Record searches by GP practices; 3) Posters and website adverts in public areas of Bristol and Cardiff; and 4) Self-referral via social media adverts through an online study recruitment agency (Lindus Health: https://www.lindushealth.com). Full details, including changes during the trial, are in the Supplementary Materials.

### Randomisation

Participants were randomised to receive either the ‘active’ CBM or ‘sham’ CBM (see below for details). Group allocation was determined using the desktop application MinimPy (Saghaei & Saghaei, 2011), and was implemented by a researcher not involved in recruitment or data analysis. Randomisation was minimised on gender, age (<35 years; ≥35 years), and depression severity (PHQ-9 score <17 vs. ≥17).

### Sample size

Our prior study indicated that active CBM, relative to sham CBM, led to increased activation to happy vs. sad faces in left amygdala (effect size d = 0.69, p<0.05) (I. S. Penton-Voak et al., 2021). A sample size of 72 would therefore provide >80% power to detect a similar effect at a 5% alpha level. We randomised 84 participants into the study allowing for ∼20% attrition during the intervention period.

### Assessments

A flowchart showing a timeline of study assessments is presented in Figure 1.

**Figure 1.**
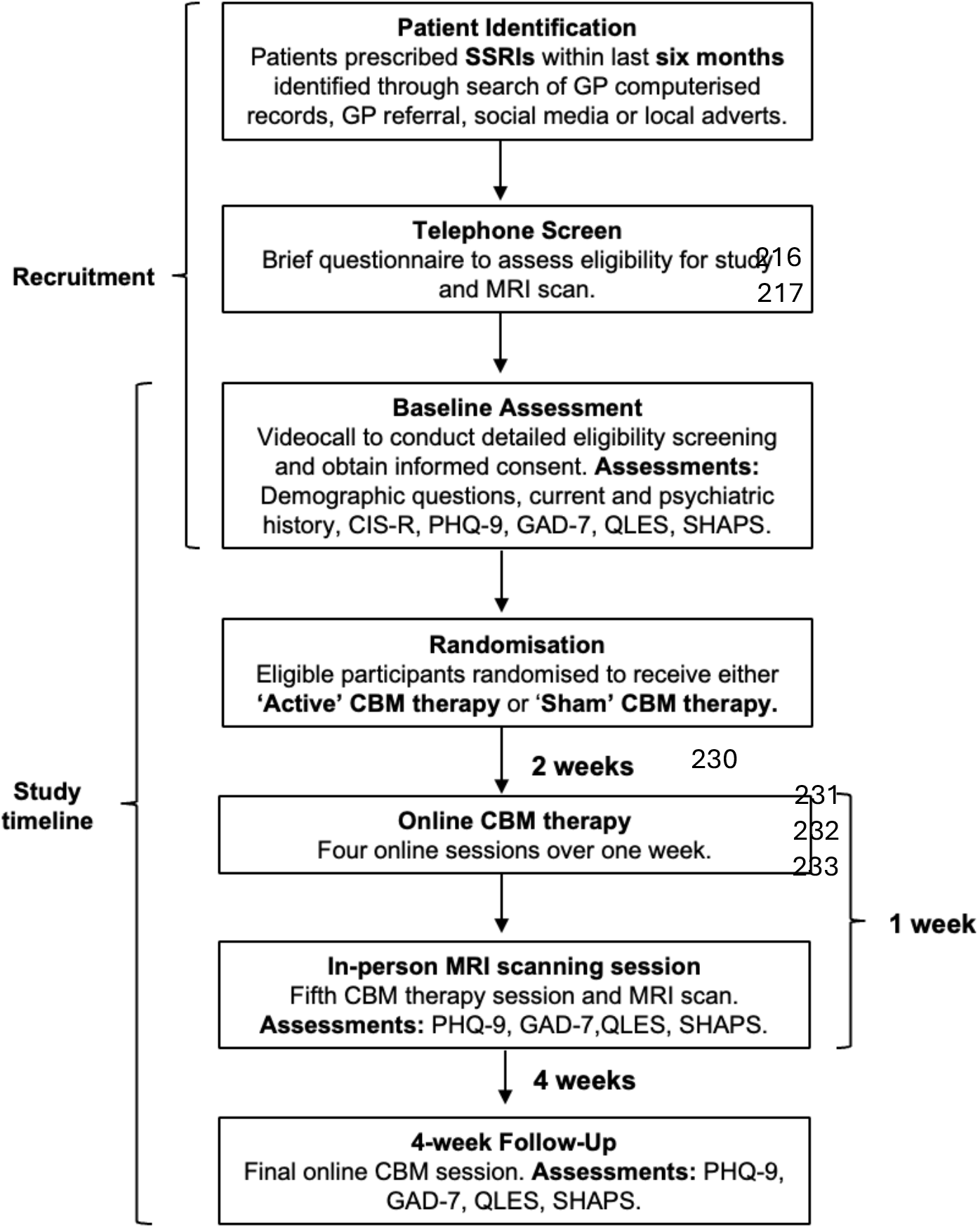
Flowchart of study assessments. Participants completed all study assessments online except for the MRI scanning session. Acronyms: SSRIs = Selective Serotonin Reuptake Inhibitors; GP = General Practitioner; MRI = Magnetic Resonance Imaging; CIS-R = Clinical Interview Schedule Revised; PHQ-9 = Patient Health Questionnaire; GAD-7 = General Anxiety Disorder questionnaire; QLES = Quality of Life Enjoyment and Satisfaction questionnaire; SHAPS = Snaith-Hamilton Pleasure Scale; CBM = Cognitive Bias Modification.

### Baseline Assessment

A videocall with a researcher lasting 60 minutes. Potentially eligible participants completed sociodemographic questions plus: ICD10 diagnosis using the Revised Clinical Interview Schedule (CIS-R)(Lewis, Pelosi, Araya, & Dunn, 1992); depressive symptoms using the Patient Health Questionnaire (PHQ-9)(Kroenke et al., 2001); anxiety symptoms using the General Anxiety Disorder questionnaire (GAD-7)(Spitzer, Kroenke, Williams, & Löwe, 2006); anhedonia using the Snaith-Hamilton Pleasure Scale (SHAPS)(Snaith et al., 1995); and the Quality of Life Enjoyment and Satisfaction Questionnaire (QLES)(Endicott, Nee, Harrison, & Blumenthal, 1993).

### Online Cognitive Bias Modification

Approximately two weeks after randomisation (to align with MRI scanner booking schedule), participants completed four computerised sessions of CBM remotely over a one-week period (see section below for details of CBM). These sessions were hosted by the Gorilla experimental platform (https://gorilla.sc), lasted 8-12 minutes each, and participants were prompted to complete the sessions if necessary. Participants completed one session per day only.

### In-person MRI scan

University Brain Research Imaging Centre. Before scanning, participants completed final safety checks, a final CBM session, and repeat online questionnaires assessing mood, anxiety, quality of life, and anhedonia (PHQ-9, GAD-7, QLES, SHAPS). Participants received instructions and practiced the three fMRI tasks on a laptop to ensure understanding. A trained radiographer conducted a 1-hour MRI scan comprising: 1) structural scan, 2) resting-state fMRI, 3) implicit facial emotion processing task, 4) working memory (n-back) task, and 5) reward learning task. Only results from the emotion processing task are reported here. Participants received £100 in Love2Shop vouchers plus travel expenses. The session lasted approximately two hours.

### 4-week follow up

Four weeks after scanning, participants completed repeat online assessments of mood, anxiety, quality of life and anhedonia (PHQ-9, GAD-7, QLES, SHAPS), plus a final baseline CBM session. Participants received £10 in Love2Shop vouchers.

### Cognitive Bias Modification task

Details of the CBM task are reported elsewhere (I. S. Penton-Voak et al., 2021) and in Supplementary Materials. CBM involved five online sessions of training accessible via phone, tablet, or computer. Sessions 1–4 included a baseline block and three training blocks and session 5 added a repeat baseline block. A final session (at 4-week follow up) included only a baseline block. The first session began with psychoeducation on cognitive biases in depression. Stimuli were composite facial expressions from the Karolinska Directed Emotional Faces dataset (Lundqvist, Flykt, & Öhman, 1998), identical to previous CBM and fMRI studies (I. S. Penton-Voak et al., 2021; I. S. Penton-Voak et al., 2012). Each face was morphed across 15 frames from unambiguously happy to unambiguously sad (Figure S1). Baseline blocks (45 trials) identified the participant’s ‘balance point’—where judgments of ambiguous expressions shifted from ‘happy’ to ‘sad’. Training blocks (31 trials) provided tailored feedback based on this balance point. In the ‘active’ CBM group, feedback was given on the two faces nearest the balance point (initially judged as sad) as ‘incorrect’, encouraging ‘happy’ categorisation. In the ‘sham’ group, feedback reinforced the original balance point.

### fMRI Facial Emotion Processing Task

The fMRI task is reported elsewhere (Godlewska, Norbury, Selvaraj, Cowen, & Harmer, 2012) and in Supplementary Materials. Briefly, the task was a simple block design measuring incidental processing of sad, happy and fearful facial expressions. Participants judged the gender of the faces via button box response.

### MRI Data Acquisition

Data were acquired using a 3T Siemens Prisma scanner (32-channel head coil). fMRI data were acquired using a multiband (or simultaneous multi-slice) 2D echo-planar imaging (EPI) sequence (MB acceleration factor = 4, integrated parallel imaging technique (iPat) = OFF, Repetition time (TR) = 1500ms; TE = 32ms; 2.0mm x 2.0mm x 2.0mm voxels; Flip angle = 70°, anterior-to-posterior acquisition, interleaved slices, 519 volumes). A structural scan was also acquired using a T1-weighted magnetization-prepared rapid gradient-echo (MPRAGE) sequence (TR = 2100ms; TE = 3.24ms; Inversion time = 850ms; flip angle = 8°; Field of View = 256mm x 256mm^2^, 1.0 x 1.0 x 1.0mm^3^ voxel size, 176 slices) for preprocessing of the EPI data.

### Statistical analyses

#### Intervention check

An intervention check was done to determine whether active CBM had the intended effect of shifting classification of emotional facial expressions (balance point). Balance points that fell 3 times above or below the interquartile range were considered outliers and were removed. Multiple regression models were conducted with group (active/sham CBM) as the predictor variable and the post-training balance points at the scanning session and 4-week follow-up as the outcome variables. Models were adjusted for the pre-training balance point and the minimisation variables (age, gender and depression severity (PHQ-9 score) at baseline). All multiple regressions were performed in R (Version 2023.06.0+421) and on all available data.

#### fMRI Behaviour

Participants that scored below chance (<50% accuracy) in the gender discrimination task were removed from analyses. A logistic regression was conducted to investigate whether gender discrimination accuracy (outcome variable) was different between active vs. sham CBM groups.

#### fMRI Analysis

##### Pre-processing

The fMRI data were pre-processed and analysed using Statistical Parametric Mapping, version 12 (SPM 12: https://www.fil.ion.ucl.ac.uk/spm/software/spm12). Structural scans were segmented into grey matter, white matter and CSF tissue classes, and normalised to Montreal Neurological Institute (MNI) space. Functional images were unwarped (using field maps) and realigned to the first image using six rigid-body realignment parameters (x, y, z, roll, pitch, yaw), slice-time corrected to the first slice in each acquisition (to account for the multiband sequence), co-registered to structural images, normalised to MNI space using tissue probability maps and forward deformation fields from the segmented structural images, and smoothed using an 6mm Full Width Half Maximum (FWHM) Gaussian kernel. Full details of motion correction of fMRI data are available in Supplementary Materials.

##### First-level modelling

In this block design, the onsets and durations for three explanatory variables of interest were modelled: ‘fearful faces’, ‘happy faces’ and ‘sad faces’. These task-related regressors were convolved with a canonical hemodynamic response function (hrf) and the denoised images were filtered using a temporal high pass filter of 128 secs in SPM. Blocks of fixation cross acted as the implicit baseline. Nuisance regressors were added including 24 movement parameters (six parameters estimated during realignment, the squares of the six realignment parameters, six parameters that were the “spin-history” of realignment (realignment from preceding volume) and the squares of the six spin-history parameters). Subject-specific outlier scans (identified during motion correction) were also added as nuisance regressors.

##### Group-level statistics

The main effects of Group (active CBM/sham CBM) and Emotion (fearful, happy and sad faces) were examined with a full factorial model, plus the Group x Emotion interaction effects with happy vs. sad faces as the primary outcome measure (H1) and happy vs. fearful faces as the secondary outcome measure (H2) (F tests). Independent-samples t tests were used to determine the direction of the effect. Unadjusted models, and models adjusted for age and gender are reported.

##### Region of Interest analyses

Primary and secondary analyses were conducted using two preregistered Regions Of Interest (ROIs): 1) bilateral amygdalae, and 2) medial prefrontal cortex (mPFC). The ROIs were selected based on an independent pilot dataset using this task (Ian S. Penton-Voak et al., 2021) and previous literature (Nord et al., 2021). Details of how the ROIs were definied are in Supplementary Materials. Effects were reported if they survived small volume correction (SVC) with an initial cluster defining threshold of p<0.001 uncorrected and voxel level threshold of p<0.05 FWE-corrected. All coordinates are reported in MNI space.

#### Exploratory analyses

To explore differences between negative emotions, the interaction effect between Group and sad vs. fearful faces was examined within ROIs and across the whole brain. Voxels surviving a voxel-level threshold of p<0.05 FWE-corrected are reported.

##### Change in balance point

To explore whether neural activity was related to a better response to CBM, participant’s change in balance point scores from a) pre-training to scanning session and b) pre-training to 4-week follow-up were added as covariate to models. The group x change in balance point interaction on neural activity was tested. All exploratory analyses used a whole-brain analysis approach (cluster defining threshold p<0.001 uncorrected, k = 20) with a cluster-level threshold of p<0.05 FWE-corrected.

##### Functional connectivity

To investigate functional connectivity between mPFC (group x happy-fearful faces interaction) and other brain regions during the task, we conducted a generalised psychophysiological interaction (PPI) analysis (McLaren, Ries, Xu, & Johnson, 2012). A 10mm sphere around the peak activation within mPFC (MNI coordinates: 14, 62, 14) was used as the seed region. Full PPI details are in Supplementary Methods. A full factorial model was used to test the main effect of group (active/sham CBM) on mPFC functional connectivity to happy, fearful and sad faces. Additional models investigated any group x change in balance point interactions on mPFC connectivity.

##### Change in depressive symptoms

To understand the clinical relevance of CBM-elicited neural effects, neural activity showing differences between CBM groups was extracted from the peak voxel (unless otherwise specified). Multiple regression models tested whether neural activity predicted depression symptoms (PHQ-9) at MRI scanning session or 4-week follow where ‘CBM group’ was the interaction term. Models were adjusted for baseline depression symptoms, age and gender. Anxiety symptoms (GAD-7), quality of life (QLES) and anhedonia (SHAPS) were also explored as outcomes.

## Results

### Participant characteristics

Eighty-four participants were randomised but two in error: a Consort Diagram is shown in Figure S2. Participant characteristics at baseline assessment are presented in Table 1. There were no differences in age, gender, depressive symptoms (PHQ-9), anxiety (GAD-7), panic (PHQ-panic), anhedonia (SHAPS) or quality of life (QLES) between active and sham CBM groups. Changes in clinical symptoms (e.g., depressive symptoms) from baseline to 4-week follow up are presented in Supplementary Materials (Figures S4-S8). More than half of participants (n = 49) were taking sertraline at baseline, whilst other SSRI medications included fluoxetine (n = 15), citalopram (n = 14) and escitalopram (n = 4). The median medication duration at baseline was 2-3 months. All participants were taking an SSRI at baseline and scanning session, and seven participants had stopped at 4-week follow-up.

**Table 1.** Participant characteristics at baseline assessment.

| Demographic variables (Mean ± SD) | Active CBM group<br>(n = 41) | Sham CBM group<br>(n = 41) |
| --- | --- | --- |
| Age | 30.95 ± 11.66 | 33.49 ± 10.00 |
| Gender (Male/Female/Other) | 12/26/3 | 13/26/2 |
| Depressive symptoms (PHQ-9) | 17.56 ± 3.67 | 17.37 ± 2.92 |
| Anxiety (GAD-7) | 13.12 ± 4.28 | 14.02 ± 4.16 |
| Panic (PHQ-panic) | 2.32 ± 2.25 | 2.07 ± 2.23 |
| Anhedonia (SHAPS) | 21.78 ± 6.93 | 22.95 ± 7.27 |

| Demographic variables (Mean ± SD) | Active CBM group<br>(n = 41) | Sham CBM group<br>(n = 41) |
| --- | --- | --- |
| Quality of life (QLES) | 36.85 ± 7.05 | 35.24 ± 7.35 |

Note: Sample includes all participants correctly randomised (n=82). PHQ-9 = Patient Health Questionnaire; GAD-7 = General Anxiety Disorder questionnaire; PHQ-panic = Patient Health Questionnaire for panic; SHAPS = Snaith-Hamilton Pleasure Scale; QLES = Quality of Life Enjoyment and Satisfaction Questionnaire.

### Intervention check

There was strong evidence that active CBM was effective at positively shifting classification of emotional facial expressions (increasing post-training balance point) at MRI scanning session (β = 2.07, 95% CI [1.20–2.94], *p*<.001, n=71) and 4-week follow-up (β = 2.32, 95% CI [1.41– 3.24], *p*<.001, n=65), adjusting for pre-training balance point, age, gender and mood at baseline (Figure 2).

**Figure 2.**
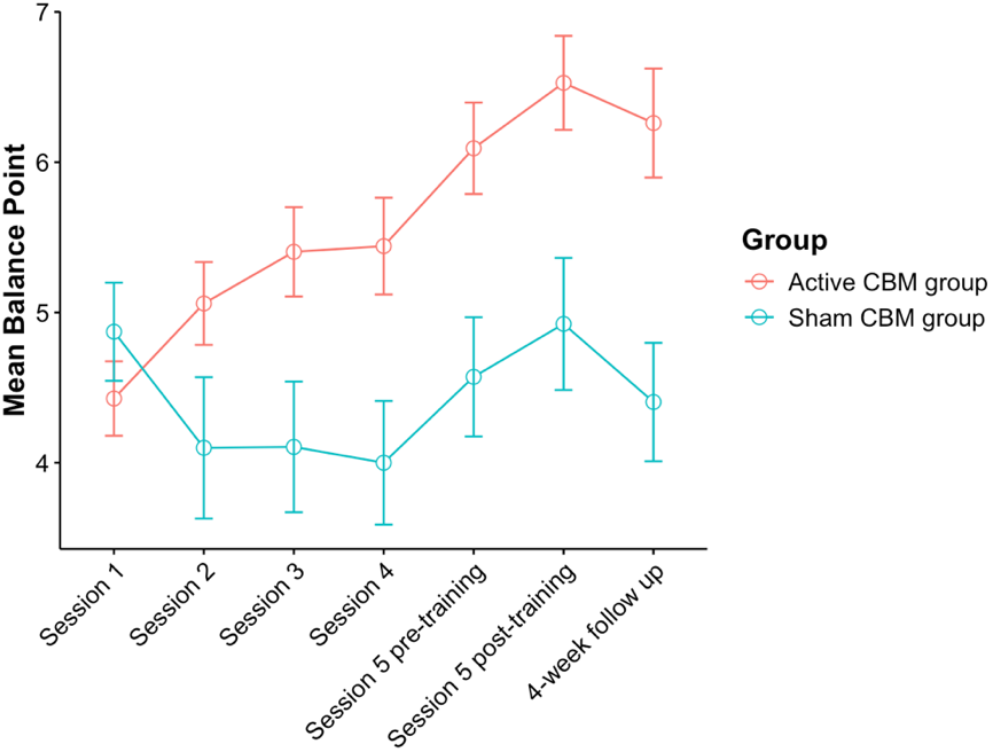
Change in balance points over time between groups. Balance points were measured at the beginning of each CBM session which occurred in the one-week period leading up to, and for Session 5 at the time of, the MRI scanning session. Error bars represent the standard error of the mean. More positive balance point relates to categorising more faces as happy rather than sad.

### fMRI data quality control

Eight participants had missing task fMRI imaging data, five had problematic and unrepairable head movement (FD > 0.3mm) and one with an incidental finding that were excluded from analysis (final sample size n=59; active CBM = 28, sham CBM = 31). fMRI task accuracy was high (median = 98%, IQR = 95% - 99%). There was no evidence that the movement parameters differed between groups (Table S1, Figure S3), or percentage accuracy on the task (*t*[1, 56.4] = 0.17, *p*=.86).

### Primary fMRI analyses

Across groups, a neural network involved with processing of emotional faces (average effect of viewing happy, fearful and sad faces compared to implicit baseline) was evoked including bilateral amygdala and mPFC regions of interest (Table S2). There was no evidence for main effects of group (active/sham CBM) or emotion (happy/fearful/sad faces) on BOLD signal in the bilateral amygdalae or mPFC. Adjusting for age and gender did not change this effect.

#### Group x Emotion Interactions

There was no evidence for a group x happy-sad contrast interaction (primary outcome measure). However, there was weak evidence for a group x happy-fear contrast interaction (secondary outcome measure) in mPFC ROI (T= 3.38, *p*=.048 small volume corrected, peak= 14, 62, 14) which was driven by reduced activity to happy faces and enhanced activity to fearful faces in the active CBM group compared to sham CBM group (Figure 3a). This interaction effect was robust to adjustment for age and gender (T= 3.36, *p*=.05 small volume corrected, peak= 14, 62, 14). There was no evidence for any group x emotion interactions in bilateral amygdalae.

**Figure 3.**
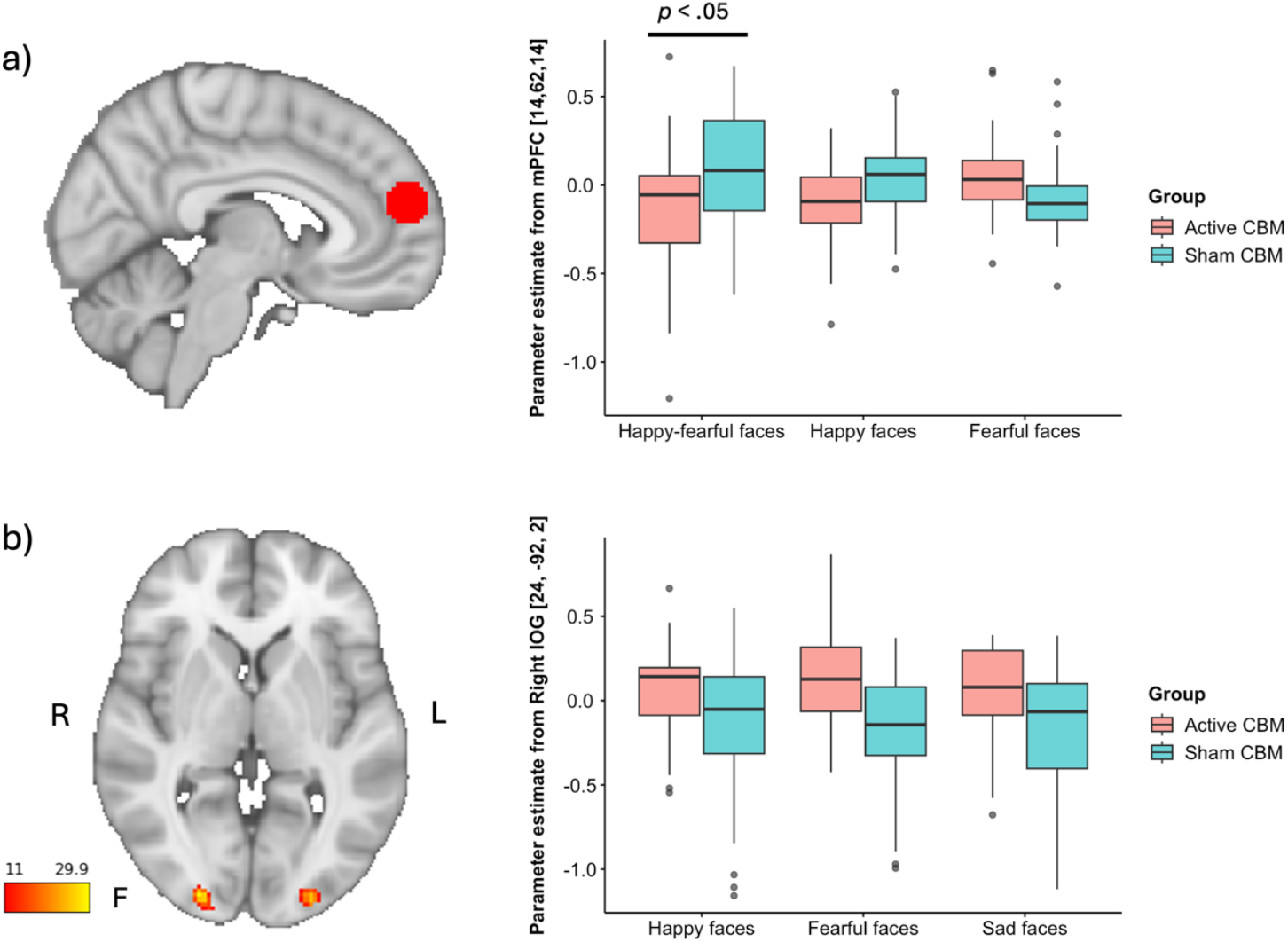
Univariate differences in neural activity between CBM groups. a) Medial Prefrontal Cortex region (binary ROI mask) overlaid onto a standard MNI brain template accompanied by a box plot of parameter estimates extracted from the peak mPFC voxel. Box plots show differences in parameter estimates between CBM groups for happy-fearful faces (group x contrast interaction) and separately for happy and fearful faces. b) Bilateral Inferior Occipital Gyrus (IOG) region overlaid onto a standard MNI brain template accompanied by a box plot of parameter estimates extracted from the peak voxel in right IOG (main effect of CBM group) across happy, fearful and sad faces. The colour bar on the left represents the F statistic.

### Pre-specified sensitivity analyses

To assess the impact of type of SSRI medication, medication duration and age on the primary fMRI findings (group x happy-fearful faces contrast interaction). Analyses restricted to participants taking Sertraline (most commonly prescribed SSRI in sample, n= 36) showed the effect in mPFC was qualitatively similar to the full sample (n=59). Differences in mPFC activity in relation to age (31-50 years vs. 19-30 years) and medication duration (0-2 months vs. 2-6 months) are presented in Figures S9 and S10.

### Exploratory fMRI analyses

#### Group x sad-fear interaction

There was no evidence for a group x sad-fear contrast interaction in mPFC or bilateral amygdalae in unadjusted and adjusted models.

#### Whole-brain analyses

There was no evidence for a main effect of CBM group in the unadjusted model. However, adjusting for age and gender revealed moderate evidence for a main effect of CBM group in right Inferior Occipital Gyrus (right IOG) (T= 5.47, *p*=.008 voxel-level FWE-corrected, peak= 24, -92, 2), and weak evidence in left IOG (T= 4.99, *p*=.06 voxel-level FWE-corrected, peak= -26, - 94, 2) where the active CBM group had enhanced activity in bilateral IOG compared to sham CBM group (Figure 3b). There was no evidence for a main effect of emotion or group x emotion contrast interaction across the whole brain.

#### Change in Balance Point

There was evidence for a group x change in balance point interaction from pre-training to 4-week follow-up (but not from pre-training to MRI scanning session) across all three emotions (happy, fearful, sad faces) in right IOG (T=4.90, 103 voxels, p=.043, peak= 34, -82, 10, cluster-level corrected) and right Inferior Parietal Cortex (right IPC) (T=4.52, 126 voxels, peak= 26, - 50, 46, cluster-level corrected) (Figure 4a&b). There was also evidence for a group x change in balance point (pre-training to 4-week follow-up) interaction specifically for happy-fear contrast in left Middle Temporal Gyrus (left MTG) (T=5.12, 91 voxels, p=.039, peak= -50, -48, 6, cluster-level corrected) (Figure 4c). For all three interaction effects, a positive shift in balance point (more happy classifications) was positively related to activity in these regions in the active CBM group, but negatively related to activity in these regions in the sham CBM group.

**Figure 4.**
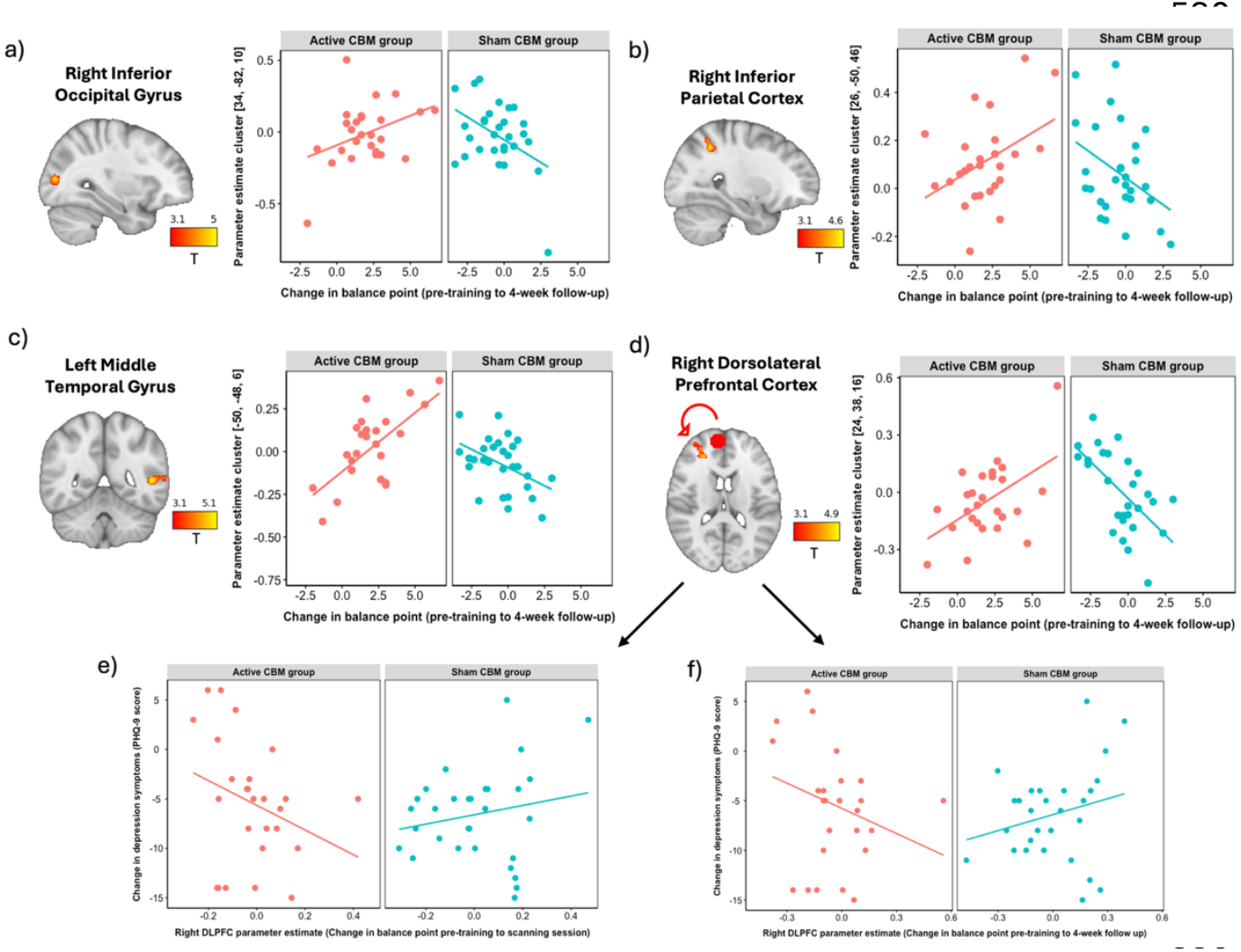
Neural activity differentially modulated by change in balance point between groups. Region of neural activity overlaid onto a standard MNI template accompanied by scatterplots, split by CBM group, showing the change in balance point (from pre-training to 4-week follow up) against parameter estimates extracted from the cluster in a) Right Inferior Occipital Gyrus (across all faces), b) Right Inferior Parietal Cortex (across all faces), c) Left Middle Temporal gyrus (happy-fearful faces contrast), and d) Right Dorsolateral Prefrontal Cortex (across all faces). Parameter estimates in Right Dorsolateral Prefrontal Cortex represent functional connectivity with mPFC (seed region) which is shown by the mPFC mask and arrow on the template brain. Bottom panel shows parameter estimates extracted from right DLPFC cluster that was associated with a change in balance point from e) pre-training to scanning session and f) pre-training to 4-week follow-up, against change in depression symptoms (PHQ-9 score) from baseline 4-week follow-up. Negative values for the change in PHQ-9 score represent a reduction in depressive symptoms. Colour bars represent the T statistic. Template brains are presented in radiological view.

#### Medial Prefrontal Cortex Functional Connectivity

There was no evidence for a main effect of CBM on mPFC functional connectivity. However, there was evidence for a group x change in balance point (from pre-training to scanning session) interaction across all three emotions in right Dorsolateral Prefrontal Cortex (right DLPFC) (T=4.54, 245 voxels, p<.001, peak= 28, 44, 14 cluster-level corrected) and left cerebellum (T=4.21, 141 voxels, p=.002, peak= -32, -68, -22 cluster-level corrected). The interaction effect in right DLPFC was also present in relation to change in balance point from pre-training to 4-week follow-up (T=4.77, 170 voxels, p=.003, peak= 24, 38, 16 cluster-level corrected) (Figure 4d). For all three interaction effects, a positive shift in balance point (more happy classifications) was positively related to mPFC-right DLPFC and mPFC-left cerebellum functional connectivity in the active compared to sham CBM group.

#### Change in Depressive Symptoms

There was no evidence that activity in mPFC (group x happy-fear contrast interaction) predicted change in depressive symptoms. However, there was weak evidence that functional connectivity between mPFC and right DLPFC (group x change in balance point interaction) was differentially predictive of change in depressive symptoms from baseline to 4-week follow-up between groups (standardised beta estimate = -0.27, 95% CI -0.54 – 0.01, *p*=.055; standardised beta estimate = -0.27, 95% CI -0.54 – 0.00, *p*=.05, respectively). Higher mPFC-DLPFC functional connectivity was related to a reduction in depressive symptoms in the active CBM group compared to sham CBM group (Figure 4e and f). There was no evidence that neural activity was related changes in symptoms of anxiety, panic, quality of life or anhedonia.

## Discussion

This study broadens our understanding of how CBM changes emotional processing in primary care patients with depression taking SSRIs. As we broadly predicted, active CBM (vs. sham) altered BOLD signal in the mPFC (but not amygdalae) in response to happy-fearful faces. Contrary to our predictions, this effect was driven by increased BOLD signal for fearful faces and decreased BOLD signal for happy faces. Additionally, active CBM increased BOLD signal in the IOG during facial processing, irrespective of emotional valence. Exploratory analyses showed that individuals that responded well to CBM (larger shif t in balance point) had increased connectivity between the mPFC and right DLPFC, and the magnitude of this connectivity change correlated with greater reduction in depressive symptoms. Overall, this suggests CBM improves negative emotional biases to ambiguous facial expressions by modulating activity relating to both attentional (high-level) and perceptual (low-level) processing of faces, where mPFC-DLPFC connectivity may play a crucial role in converting these perceptual changes into mood improvements.

Our pre-registered hypotheses that active CBM would increase BOLD response to positive emotions (happy-sad and happy-fear contrasts) in the amygdalae and mPFC were partially supported. Although we observed mPFC modulation, there was no evidence for changes in amygdalae. The neural locus of CBM’s effects was not unexpected given prior evidence (Fu et al., 2004; Wiers & Wiers, 2017) and our earlier imaging work in individuals with depression who were not taking antidepressants (I. S. Penton-Voak et al., 2021). However, a recent meta-analysis reported that psychotherapies (including CBT, mindfulness and other cognitive therapies) changed responses to negative information by acting on the mPFC while antidepressants acted on amygdalae. This suggests a division of mehcnistic processes with psychotherapies targeting cognitive processes and dysfunctional schemata at the cortical level (Gotlib & Joormann, 2010) and antidepressants modulating subcortical threat-detection circuits within the amygdalae (LeDoux, 2003). However, since all participants were taking medication, CBM’s potential effects on the amygdalae may have been masked, nullified or overwritten by antidepressants. Further studies randomising participants to SSRI or placebo (alongside CBM) are needed to resolve these questions.

A novel feature of this work is the direction of BOLD signal changes in the mPFC induced by CBM, which contrasts with our previous work in a sample not taking antidepressants showing reduced mPFC activity to happy faces (I. S. Penton-Voak et al., 2021).

Again, this discrepancy may be due to SSRI administration. Currently, no neurocognitive model reliably predicts when psychological interventions will increase or decrease BOLD signal in the frontal cortex. Psychotherapy has been reported to produce both outcomes, perhaps due to heterogeneity in clinical diagnosis, medication, comorbidity, and therapy type between studies (Marwood, Wise, Perkins, & Cleare, 2018; Yoshimura et al., 2014). This highlights the need for a richer neurocognitive model of how psychological therapies affect prefrontal function, particularly given CBM can modulate neuronal alpha oscillations (Martínez-Maldonado et al., 2020) which are negatively correlated with BOLD signal (Scheeringa, Mazaheri, Bojak, Norris, & Kleinschmidt, 2011). Resolving this will require multimodal studies such as simultaneous fMRI-EEG recordings.

Additionally, the direction of mPFC activity may be less important than its connectivity with other brain regions which varied between participants. A greater shift in balance point was associated with greater mPFC-DLPFC functional connectivity which predicted improvement in depression symptoms four weeks later. This suggests mPFC-DLPFC connectivity may be crucial for CBM’s clinical effects. Participants who showed the largest changes in emotional categorisations during CBM showed changes in their mPFC-DLPFC connectivity and reduced their depressive symptomatology. This would be consistent with neuromodulation interventions (e.g., Transcranial Magnetic Stimulation; TMS) that improve top-down control of affective information as the DLPFC is a key region targeted in depression (Kan et al., 2023; Nejati, Majdi, Salehinejad, & Nitsche, 2021). However, this causal pathway cannot be confirmed from these results. In addition to top-down modulation in mPFC, increased BOLD signal was also observed in IOG, implicated in processing lower-level features (Kim, Nanavaty, Ahmed, Mathur, & Anderson, 2021). While CBM has been proposed to resolve high-level (e.g., attentional) biases, here there is evidence it acted on both high- and low-level (e.g., perceptual) biases (Roiser et al., 2012). Similar modulation has been observed in response to CBT (Ren et al., 2025) demonstrating CBM’s capacity to exert effects across the cortical hierarchy.

Some neural mechanisms underpinning CBM were related to participants’ responsiveness to CBM (i.e., individual differences in the change in balance point from baseline). In the active CBM group, a greater increase in balance point (reflecting a less negative bias in interpreting ambiguous faces) was associated with enhanced BOLD signal in right IOG, right IPC and mPFC-right DLPFC functional connectivity irrespective of emotional valence. Balance point also modulated the left MTG in the active CBM group, specifically for happy-fearful faces. The IOG and IPC have been implicated in processing visual features and representing the motivational salience of emotions (Kim et al., 2021). Whereas the MTG supports the recognition and social processing of faces (Yun et al., 2017). Interestingly, a previous study showed individuals with depression had decreased response in left MTG to happy-fearful faces suggesting CBM may reverse this effect and enhance positive biases to happy faces (Kustubayeva, Eliassen, Matthews, & Nelson, 2023; Ian S. Penton-Voak et al., 2017). This supports the idea that CBM might increase awareness and recognition of emotional faces (particularly happy and fearful).

Interestingly, we observed no changes in activity to sad faces despite participants being trained on happy and sad faces during CBM. Again, this may reflect intact recognition of this emotion in both groups (Dalili, Penton-Voak, Harmer, & Munafò, 2015) or the influence of SSRI medication, which was common to both groups. Alternatively, participants may have already desensitised to sad faces and reacted more strongly to fearful faces which were novel (although all fMRI stimuli were different to CBM stimuli). Indeed, training on low-arousal emotions (e.g., happy-sad) has been shown to generalise to high-arousal emotions (e.g., happy-angry) (Dalili et al., 2017). Nevertheless, our findings on individual balance points are concordant with our meta-analysis of behavioural CBM studies that showed mood improvements were mediated by the post-training balance point (Rumeysa Kuruoğlu, Suddell, & Penton-Voak, 2025). Importantly, the strongest effects were observed for the change in balance point from pre-training to 4-week follow-up (rather than immediately after CBM). This suggests robust changes in emotion perception are important and supports neurocognitive models proposing that repeated emotional exposure through social interactions is needed to change negative emotional biases over time (Harmer et al., 2009). Future studies are therefore needed to maximise responsiveness to CBM by identifying predictors (e.g., motivation) (Gladwin, Wiers, & Wiers, 2016) and improving engagement (e.g., through gamification) (R. Kuruoğlu et al., 2025).

### Strengths and Limitations

A key strength of this study is that it was conducted in a clinical sample of participants with depression recruited from primary care. The study used a double-blind randomised controlled trial design and the protocol and data analysis plan were pre-registered. The fMRI analyses were adequately powered to detect pre-specified differences in BOLD signal between groups. The limitations of the study are that only one fMRI scan was conducted after CBM so baseline neural activity (before CBM), and neural changes in response to CBM, were not measured. Medication dose was uncontrolled and could not be added as a covariate due to different medications being used in the sample, however all were using SSRIs that have similar serotonergic mechanisms of action. Finally, the study was not powered to detect changes in clinical symptoms and so although the evidence for the effects on depressive symptoms and panic are promising, these findings should be treated cautiously.

### Clinical Implications

CBM modified neural mechanisms associated with negative emotional biases in primary care patients with depression who were taking SSRI antidepressants. Participants receiving active CBM showed enhanced prefrontal cortex connectivity that was associated with greater improvements in depressive symptoms. CBM has potential as an adjunctive treatment to pharmacotherapy. It could prove beneficial to those facing barriers to psychotherapy such as long waiting lists, treatment costs, or stigma (Titov et al., 2010). Further research is therefore needed to assess CBM’s clinical and cost effectiveness, the number of sessions required and the duration of effect.

## Conclusions

The findings suggest that CBM modified brain circuits involved in emotional perception in a clinical sample of patients with depression recruited from primary care who were taking SSRIs. The neural locus of these effects is similar to other psychotherapies (e.g., CBT) but distinctive, and complimentary, to SSRIs. Crucially, heterogeneity in behavioural and clinical response to CBM may stem from differences in cortical response, particularly mPFC-DLPFC connectivity. Thus, CBM may be a low-cost, scalable way of changing emotional processing circuits in depression and is a promising adjunct therapy to antidepressants.

## Data Availability

The data and code that support the findings of this study are currently available to peer reviewers to view on the Open Science Framework: https://osf.io/rxqy4/overview?view_only=842fb8762c74467cba363848887fdfca and will be made openly available on the University of Bristol Research Data Storage Facility after manuscript acceptance.

## Acknowledgements

We would like to acknowledge Lindus Health for their assistance in recruiting participants for this study. We would like to thank the participants that took part and the GP practices who supported the study. We would like to thank the trial steering committee that oversaw this trial and provided useful feedback and suggestions. Chat GPT was used to reduce the length of the main text in this publication. AI was used for no other purposes.

## Ethical Standards

The authors assert that all procedures contributing to this work comply with the ethical standards of the relevant national and institutional committees on human experimentation and with the Helsinki Declaration of 1975, as revised in 2011. The trial was conducted according to the Athens 1996 ICH Guidelines for Good Clinical Practise E6 (R2). All participants provided informed consent beforehand and were compensated for their time.

## Competing Interests

MM & IPV receive royalties from Cambridge Cognition, through the University of Bristol, for software for the assessment of emotion recognition.

## Author Contributions

**Charlotte Crisp**: conceptualisation, formal analysis, investigation, methodology, project administration, software, supervision, visualisation, preparation of original draft. **Sean Fallon**: conceptualisation, funding acquisition, formal analysis, methodology, software, supervision, review and editing. **Alison Burns**: data curation, investigation, project administration, resources, review and editing. **Rumeysa Kuruoglu**: investigation, review and editing. **Jennifer Ferrar**: investigation, project administration, review and editing. **Nicola Wiles**: conceptualisation, funding acquisition, resources, review and editing. **David Kessler**: conceptualisation, funding acquisition, resources, review and editing. **Marcus Munafo**: conceptualisation, funding acquisition, review and editing. **Ian Penton-Voak**: conceptualisation, funding acquisition, methodology, resources, software, supervision, review and editing.

## Notes

**Financial Support** This work was supported by an experimental medicine challenge grant awarded by the Medical Research Council (MR/S035648/1) to Professor Ian Penton-Voak. This study has been delivered through the National Institute for Health and Care Research (NIHR) Bristol Biomedical Research Centre (BRC). The views expressed are those of the author(s) and not necessarily those of the MRC, the NIHR or the Department of Health and Social Care.

### Clinical Trial

ISRCTN37448835

### Clinical Protocols

https://doi.org/10.17605/OSF.IO/FYUJN

### Funding Statement

This work was supported by an experimental medicine challenge grant awarded by the Medical Research Council (MR/S035648/1) to Professor Ian Penton-Voak. This study has been delivered through the National Institute for Health and Care Research (NIHR) Bristol Biomedical Research Centre (BRC). The views expressed are those of the author(s) and not necessarily those of the MRC, the NIHR or the Department of Health and Social Care.

### Author Declarations

Ethics approval was obtained from the NHS Health and Care Research Wales (REC reference number 20/LO/1118). Health Research Authority (HRA) approval was also obtained.

## References

Al-Harbi, K. S. (2012). Treatment-resistant depression: therapeutic trends, challenges, and future directions. Patient Prefer Adherence, 6, 369–388. doi:10.2147/ppa.S29716

Bains, N., & Abdijadid, S. (2025). Major Depressive Disorder. In StatPearls. Treasure Island (FL) ineligible companies. Disclosure: Sara Abdijadid declares no relevant financial relationships with ineligible companies.: StatPearls Publishing

Copyright © 2025, StatPearls Publishing LLC.

Chan, S. W. Y., Goodwin, G. M., & Harmer, C. J. (2007). Highly neurotic never-depressed students have negative biases in information processing. Psychological Medicine, 37(9), 1281–1291. doi:10.1017/s0033291707000669

Dalili, M. N., Penton-Voak, I. S., Harmer, C. J., & Munafò, M. R. (2015). Meta-analysis of emotion recognition deficits in major depressive disorder. Psychol Med, 45(6), 1135–1144. doi:10.1017/s0033291714002591

Dalili, M. N., Schofield-Toloza, L., Munafò, M. R., & Penton-Voak, I. S. (2017). Emotion recognition training using composite faces generalises across identities but not all emotions. Cogn Emot, 31(5), 858–867. doi:10.1080/02699931.2016.1169999

Disner, S. G., Beevers, C. G., Haigh, E. A. P., & Beck, A. T. (2011). Neural mechanisms of the cognitive model of depression. Nature Reviews Neuroscience, 12(8), 467–477. doi:10.1038/nrn3027

Endicott, J., Nee, J., Harrison, W., & Blumenthal, R. (1993). Quality of Life Enjoyment and Satisfaction Questionnaire: a new measure. Psychopharmacol Bull, 29(2), 321–326.

Fu, C. H. Y., Williams, S. C. R., Cleare, A. J., Brammer, M. J., Walsh, N. D., Kim, J., … Bullmore, E. T. (2004). Attenuation of the neural response to sad faces in major depression by antidepressant treatment - A prospective, event-related functional magnetic resonance imaging study. Archives of General Psychiatry, 61(9), 877–889. doi:10.1001/archpsyc.61.9.877

Gladwin, T. E., Wiers, C. E., & Wiers, R. W. (2016). Chapter 15 - Cognitive neuroscience of cognitive retraining for addiction medicine: From mediating mechanisms to questions of efficacy. In H. Ekhtiari & M. P. Paulus (Eds.), Progress in Brain Research (Vol. 224, pp. 323–344): Elsevier.

Godlewska, B. R., Norbury, R., Selvaraj, S., Cowen, P. J., & Harmer, C. J. (2012). Short-term SSRI treatment normalises amygdala hyperactivity in depressed patients. Psychological Medicine, 42(12), 2609–2617. doi:10.1017/S0033291712000591

Gotlib, I. H., & Joormann, J. (2010). Cognition and depression: current status and future directions. Annu Rev Clin Psychol, 6, 285–312. doi:10.1146/annurev.clinpsy.121208.131305

Hallquist, M. N., Hwang, K., & Luna, B. (2013). The nuisance of nuisance regression: spectral misspecification in a common approach to resting-state fMRI preprocessing reintroduces noise and obscures functional connectivity. Neuroimage, 82, 208–225. doi:10.1016/j.neuroimage.2013.05.116

Harmer, C. J., Goodwin, G. M., & Cowen, P. J. (2009). Why do antidepressants take so long to work? A cognitive neuropsychological model of antidepressant drug action. Br J Psychiatry, 195(2), 102–108. doi:10.1192/bjp.bp.108.051193

Horne, C. M., Marr-Phillips, S. D. M., Jawaid, R., Gibson, E. L., & Norbury, R. (2017). Negative emotional biases in late chronotypes. Biological Rhythm Research, 48(1), 151–155. doi:10.1080/09291016.2016.1236461

Kan, R. L. D., Padberg, F., Giron, C. G., Lin, T. T. Z., Zhang, B. B. B., Brunoni, A. R., & Kranz, G. S. (2023). Effects of repetitive transcranial magnetic stimulation of the left dorsolateral prefrontal cortex on symptom domains in neuropsychiatric disorders: a systematic review and cross-diagnostic meta-analysis. The Lancet Psychiatry, 10(4), 252–259. doi:10.1016/S2215-0366(23)00026-3

Kim, H., Nanavaty, N., Ahmed, H., Mathur, V. A., & Anderson, B. A. (2021). Motivational Salience Guides Attention to Valuable and Threatening Stimuli: Evidence from Behavior and Functional Magnetic Resonance Imaging. Journal of Cognitive Neuroscience, 33(12), 2440–2460. doi:10.1162/jocn_a_01769

Kroenke, K., Spitzer, R., & Williams, J. (2001). The PHQ-9: validity of a brief depression severity measure. Journal of General Internal Medicine, 16(9), 606–613.

Kuruoglu, R., Attwood, A., & Penton-Voak, I. (2025). Testing the Effectiveness of a Gamified Emotional Cognitive Bias Modification Task as an Intervention for Low Mood: Randomized Controlled Trial. JMIR Serious Games, 13, e65103. doi:10.2196/65103

Kuruoglu, R., Suddell, S., & Penton-Voak, I. S. (2025). Meta-analysis of studies testing cognitive bias modification (CBM) intervention for emotion recognition bias. Current Psychology. doi:10.1007/s12144-025-08280-2

Kustubayeva, A., Eliassen, J., Matthews, G., & Nelson, E. (2023). FMRI study of implicit emotional face processing in patients with MDD with melancholic subtype. Front Hum Neurosci, 17, 1029789. doi:10.3389/fnhum.2023.1029789

LeDoux, J. (2003). The Emotional Brain, Fear, and the Amygdala. Cellular and Molecular Neurobiology, 23(4), 727–738. doi:10.1023/A:1025048802629

Lewis, G., Pelosi, A., Araya, R., & Dunn, G. (1992). Measuring psychiatric disorder in the community: a standardized assessment for use by lay interviewers. Psychol Med, 22(2), 465–486.

Li, J., Xu, C., Cao, X., Gao, Q., Wang, Y., Wang, Y., … Zhang, K. (2013). Abnormal activation of the occipital lobes during emotion picture processing in major depressive disorder patients. Neural Regen Res, 8(18), 1693–1701. doi:10.3969/j.issn.1673-5374.2013.18.007

Lundqvist, D., Flykt, A., & Öhman, A. (1998). Karolinska directed emotional faces. Cognition and Emotion.

Ma, Y. (2015). Neuropsychological mechanism underlying antidepressant effect: a systematic meta-analysis. Mol Psychiatry, 20(3), 311–319. doi:10.1038/mp.2014.24

Martfnez-Maldonado, A., Jurado-Barba, R., Sion, A., Domfnguez-Centeno, I., Castillo-Parra, G., Prieto-Montalvo, J., & Rubio, G. (2020). Brain functional connectivity after cognitive-bias modification and behavioral changes in abstinent alcohol-use disorder patients. Int J Psychophysiol, 154, 46–58. doi:10.1016/j.ijpsycho.2019.10.004

Marwood, L., Wise, T., Perkins, A. M., & Cleare, A. J. (2018). Meta-analyses of the neural mechanisms and predictors of response to psychotherapy in depression and anxiety. Neurosci Biobehav Rev, 95, 61–72. doi:10.1016/j.neubiorev.2018.09.022

McLaren, D. G., Ries, M. L., Xu, G., & Johnson, S. C. (2012). A generalized form of context-dependent psychophysiological interactions (gPPI): a comparison to standard approaches. Neuroimage, 61(4), 1277–1286. doi:10.1016/j.neuroimage.2012.03.068

McMakin, D. L., Olino, T. M., Porta, G., Dietz, L. J., Emslie, G., Clarke, G., Brent, D. A. (2012). Anhedonia predicts poorer recovery among youth with selective serotonin reuptake inhibitor treatment-resistant depression. J Am Acad Child Adolesc Psychiatry, 51(4), 404–411. doi:10.1016/j.jaac.2012.01.011

Morfini, F., Whitfield-Gabrieli, S., & Nieto-Castañón, A. (2023). Functional connectivity MRI quality control procedures in C0NN. Front Neurosci, 17, 1092125. doi:10.3389/fnins.2023.1092125

Nejati, V., Majdi, R., Salehinejad, M. A., & Nitsche, M. A. (2021). The role of dorsolateral and ventromedial prefrontal cortex in the processing of emotional dimensions. Scientiic Reports, 11(1), 1971. doi:10.1038/s41598-021-81454-7

Nord, C. L., Barrett, L. F., Lindquist, K. A., Ma, Y., Marwood, L., Satpute, A. B., & Dalgleish, T. (2021). Neural effects of antidepressant medication and psychological treatments: a quantitative synthesis across three meta-analyses. Br J Psychiatry, 219(4), 546–550. doi:10.1192/bjp.2021.16

Penton-Voak, I. S., Adams, S., Button, K. S., Fluharty, M., Dalili, M., Browning, M., … Munafò, M. R. (2021). Emotional recognition training modifies neural response to emotional faces but does not improve mood in healthy volunteers with high levels of depressive symptoms. Psychol Med, 51(7), 1211–1219. doi:10.1017/s0033291719004124

Penton-Voak, I. S., Adams, S., Button, K. S., Fluharty, M., Dalili, M., Browning, M., … Munafò, M. R. (2021). Emotional recognition training modifies neural response to emotional faces but does not improve mood in healthy volunteers with high levels of depressive symptoms. Psychological Medicine, 51(7), 1211–1219. doi:10.1017/S0033291719004124

Penton-Voak, I. S., Bate, H., Lewis, G., & Munafò, M. R. (2012). Effects of emotion perception training on mood in undergraduate students: randomised controlled trial. Br J Psychiatry, 201(1), 71–72. doi:10.1192/bjp.bp.111.107086

Penton-Voak, I. S., Munafò, M. R., & Looi, C. Y. (2017). Biased Facial-Emotion Perception in Mental Health Disorders: A Possible Target for Psychological Intervention? Current Directions in Psychological Science, 26(3), 294–301. doi:10.1177/0963721417704405

Peters, S. E., Lumsden, J., Peh, O. H., Penton-Voak, I. S., Munafò, M. R., & Robinson, O. J. (2017). Cognitive bias modification for facial interpretation: a randomized controlled trial of transfer to self-report and cognitive measures in a healthy sample. R Soc Open Sci, 4(12), 170681. doi:10.1098/rsos.170681

Power, J. D., Barnes, K. A., Snyder, A. Z., Schlaggar, B. L., & Petersen, S. E. (2012). Spurious but systematic correlations in functional connectivity MRI networks arise from subject motion. NeuroImage, 59(3), 2142–2154. doi:10.1016/j.neuroimage.2011.10.018

Power, J. D., Mitra, A., Laumann, T. O., Snyder, A. Z., Schlaggar, B. L., & Petersen, S. E. (2014). Methods to detect, characterize, and remove motion artifact in resting state fMRI. Neuroimage, 84, 320–341. doi:10.1016/j.neuroimage.2013.08.048

Ren, J., Ma, L., Wu, W., Qiu, J., Zhang, Z., Hong, Y., … Li, X. (2025). Activation Likelihood Estimation Meta-Analysis of the Effects of Cognitive Behavioral Therapy on Brain Activation in the Treatment of Depression and Anxiety Disorders. Depress Anxiety, 2025, 3557367. doi:10.1155/da/3557367

Roiser, J. P., Elliott, R., & Sahakian, B. J. (2012). Cognitive mechanisms of treatment in depression. Neuropsychopharmacology, 37(1), 117–136. doi:10.1038/npp.2011.183

Ruhe, H. G., Mocking, R. J. T., Figueroa, C. A., Seeverens, P. W. J., Ikani, N., Tyborowska, A., Schene, A. H. (2019). Emotional Biases and Recurrence in Major Depressive Disorder. Results of 2.5 Years Follow-Up of Drug-Free Cohort Vulnerable for Recurrence. Front Psychiatry, 10, 145. doi:10.3389/fpsyt.2019.00145

Saghaei, M., & Saghaei, S. (2011). Implementation of an open-source customizable minimization program for allocation of patients to parallel groups in clinical trials. Journal of Biomedical Science and Engineering, 4, 734–739. doi:10.4236/jbise.2011.411090

Scheeringa, R., Mazaheri, A., Bojak, I., Norris, D. G., & Kleinschmidt, A. (2011). Modulation of visually evoked cortical FMRI responses by phase of ongoing occipital alpha oscillations. J Neurosci, 31(10), 3813–3820. doi:10.1523/jneurosci.4697-10.2011

Snaith, R. P., Hamilton, M., Morley, S., Humayan, A., Hargreaves, D., & Trigwell, P. (1995). A scale for the assessment of hedonic tone the Snaith-Hamilton Pleasure Scale. Br J Psychiatry, 167(1), 99–103. doi:10.1192/bjp.167.1.99

Spitzer, R., Kroenke, K., Williams, J., & Löwe, B. (2006). A brief measure for assessing generalized anxiety disorder: the GAD-7. Archives of Internal Medicine, 166(10), 1092–1097.

Suddell, S., Müller-Glodde, M., Lumsden, J., Looi, C. Y., Granger, K., Barnett, J. H., … Penton-Voak, I. S. (2021). Emotional bias training as a treatment for anxiety and depression: evidence from experimental medicine studies in healthy and medicated samples. Psychol Med, 1–10. doi:10.1017/s0033291721002014

Titov, N., Andrews, G., Davies, M., McIntyre, K., Robinson, E., & Solley, K. (2010). Internet treatment for depression: a randomized controlled trial comparing clinician vs. technician assistance. PLoS One, 5(6), e10939. doi:10.1371/journal.pone.0010939

Van Meter, A., Stoddard, J., Penton-Voak, I., & Munafò, M. R. (2021). Interpretation bias training for bipolar disorder: A randomized controlled trial. J Affect Disord, 282, 876–884. doi:10.1016/j.jad.2020.12.162

Victor, T. A., Furey, M. L., Fromm, S. J., Ohman, A., & Drevets, W. C. (2010). Relationship between amygdala responses to masked faces and mood state and treatment in major depressive disorder. Arch Gen Psychiatry, 67(11), 1128–1138. doi:10.1001/archgenpsychiatry.2010.144

Warren, M. B., Pringle, A., & Harmer, C. J. (2015). A neurocognitive model for understanding treatment action in depression. Philosophical transactions of the Royal Society of London. Series B, Biological sciences, 370(1677), 20140213–20140213. doi:10.1098/rstb.2014.0213

Whitfield-Gabrieli, S., & Nieto-Castanon, A. (2012). Conn: a functional connectivity toolbox for correlated and anticorrelated brain networks. Brain Connect, 2(3), 125–141. doi:10.1089/brain.2012.0073

Wiers, C. E., & Wiers, R. W. (2017). Imaging the neural effects of cognitive bias modification training. NeuroImage, 151, 81–91. doi:10.1016/j.neuroimage.2016.07.041

Yoshimura, S., Okamoto, Y., Onoda, K., Matsunaga, M., Okada, G., Kunisato, Y., … Yamawaki, S. (2014). Cognitive behavioral therapy for depression changes medial prefrontal and ventral anterior cingulate cortex activity associated with self-referential processing. Soc Cogn Affect Neurosci, 9(4), 487–493. doi:10.1093/scan/nst009

Yun, J.-Y., Kim, J.-C., Ku, J., Shin, J.-E., Kim, J.-J., & Choi, S.-H. (2017). The left middle temporal gyrus in the middle of an impaired social-affective communication network in social anxiety disorder. Journal of Affective Disorders, 214, 53–59. doi:10.1016/j.jad.2017.01.043

Zhang, L., Yu, F., Hu, Q., Qiao, Y., Xuan, R., Ji, G., Wang, K. (2020). Effects of SSRI Antidepressants on Attentional Bias toward Emotional Scenes in First-Episode Depressive Patients: Evidence from an Eye-Tracking Study. Psychiatry Investig, 17(9), 871–879. doi:10.30773/pi.2019.0345

