## Supplementary for "Cognitive bias modification for emotional facial expressions modifies neural mechanisms in individuals taking antidepressant medication: a Randomised Controlled Trial"

**Supplementary Materials and Methods**

**Inclusion criteria**

- Aged 18-55 years.
- A new or first episode of depression (defined as being prescribed an antidepressant within six months prior to randomisation but not in the preceding six months).
- Prescribed a course of SSRI antidepressant medication (including sertraline, citalopram, dapoxetine, escitalopram, fluoxetine, fluvoxamine, paroxetine, zimelidine and vortioxetine).
- Low mood (score > 10 on Patient Health Questionnaire; PHQ-9).

Originally our inclusion criteria were defined as being prescribed an antidepressant within two weeks prior to randomisation. However, due to the difficulty in identifying and randomising participants within this narrow time frame, this criterion was relaxed to six months.

**Exclusion criteria**

- Prescribed an antidepressant for longer than six months.
- Self-reported alcohol or substance dependency, Bipolar Disorder, Schizophrenia or Psychosis, or Dementia.
- Under psychiatric care (including those referred but not yet seen) for depression.
- Unable to access online CBM sessions (via PC, laptop, smartphone, tablet).
- Could not complete questionnaires unaided or would require an interpreter.
- Taking part in another trial involving a psychological/drug therapy.
- Receiving a course of high intensity psychological therapy for depression or anxiety in the last six months.
- Had a contraindication for MRI scanning, including:
- Metal objects in or around the body which could not be removed (braces, pacemaker, metal fragments, hearing devices, accidents involving metal fragments).
- Significant hearing impairment (aids cannot be worn in the scanner).
- Significant visual impairment that could not be corrected by glasses/contact lenses e.g., double vision or loss of vision in one eye, severe cataracts.
- History of established Central Nervous System disease or injury (e.g., cerebro-vascular disease, Multiple Sclerosis, Parkinson’s Disease, Traumatic Brain Injury).
- Epilepsy, type 1 diabetes (insulin pump or electronic device attached) or thermoregulatory problems including Raynaud’s disease.
- Location-sensitive tattoos on the head, neck or genital area (participants with tattoos covering >5% of the body or longer than 20cm or multiple tattoos <20cm apart were discussed with the radiographer due to the small chance of heating during MRI scanning).
- Body Mass Index (BMI) >35 kg/m^2^.
- Too physically unwell to tolerate an MRI scan including musculo-skeletal disorders which make lying supine and still difficult.
- Claustrophobia.
- Pregnant or trying-to-become pregnant.

**Recruitment streams**

Method 1: in-consultation

General Practitioners (GPs) (Participant Identification Centres) identified patients during face-to-face in-person consultations or via videocall or telephone consultations who were starting or had very recently started (within the last six months) on an SSRI antidepressant that they thought might be suitable for the trial. They introduced the trial and asked the patient for their permission to be contacted by the research team. Verbal permission to be contacted could be taken during telephone and videocall consultations. GP practices were set in the surrounding areas of Bristol, North Somerset, South Gloucestershire, Bath and North East Somerset, Gloucestershire, Somerset and Cardiff that acted as Participant Identification Centres.

Method 2: record search

GP practices (Participant Identification Centres) conducted a search of their computerised records for potentially eligible patients (defined as those who are aged 18-55 years old, who have been recently prescribed an SSRI antidepressant – within six months). Practices were asked to exclude those who would be unsuitable due to the exclusion criteria. The record search was conducted using a combination of primary care diagnostic codes, and manual screening of resulting lists by practice staff including a practice GP. Potentially eligible patients were then either texted/ called by the GP or mailed an invitation to participate by the GP practice, asking for their permission to be contacted by the research team. Patients that did not respond after one week were sent one reminder letter/text or a follow up call by the practice. Patients were able to respond anonymously if they wished to decline participation and were able to provide a reason for declining. We also asked practices to provide anonymised data for all patients identified by the record search (age, gender, reason for exclusion by practice). This data was used to report the generalisability of results.

Methods 3 and 4: Advertisements outside the NHS

The study was also advertised via social media (e.g., twitter, websites and Lindus Health, a commercial recruitment agency), posters or equivalent materials. This method provided information about how an individual could obtain further information/ take part, including relevant contact details. Interested individuals were directed to complete an Expression Of Interest (EOI) form online using Jisc Online Surveys - an online survey tool designed for academic research (<https://www.onlinesurveys.ac.uk>) or via paper equivalent if required/preferred. The data is secure and strict information security standards were followed, and the data was processed in compliance with General Data Protection Regulations (GDPR). This included patient contact details, details of their GP and brief questions to ascertain eligibility. As part of this EOI, the potential participant was asked to confirm their agreement that the research team could inform their GP about study participation and that their GP could be contacted if the team had any concerns around issues relating to their safety (in line with the study’s policy for managing risk). In response to their EOI the research team texted/emailed suitable individuals a Participant Information Sheet (PIS) to read before the researcher called them. Those identified via Lindus Health’s social media strategy were directed to complete a pre-screener questionnaire on their website, which identified potentially suitable participants. This is similar to the EOI mentioned above. With the participant’s consent, suitable contact details were emailed to the research team.

Our preregistration specified streams 1 and 2. However, disruption to primary care in the UK during and following the Covid-19 pandemic led us to broaden our recruitment method, and most participants were recruited from the general population (recruitment streams 3 and 4) rather than primary care (streams 1 and 2). Potential participants who agreed to be contacted via any of the three recruitment methods (in-consultation/ record search/ advertising) were telephoned by the local research team and screened for suitability at the baseline appointment. Participants completed an Expression of Interest (EOI) form, including screening questions, before undergoing a screening call with a researcher to assess their initial eligibility for the study. Potentially eligible participants then took part in an online video call to do the baseline assessment with a researcher. The researcher answered any questions the participant had, confirmed initial inclusion criteria and delivered the Patient Health Questionnaire (PHQ-9) (1). Participants that scored >10 on the PHQ-9 were eligible for the study. Online written informed consent was obtained from every participant before they were randomised into the study.

**Cognitive Bias Modification task**

This Cognitive Bias Modification (CBM) task targets the recognition of emotions in facial expressions, with the aim of increasing the likelihood of making positive attributions to ambiguous emotional stimuli. Here, CBM consisted of six sessions. Sessions 1-4 each consisted of a baseline block and three training blocks which were completed over one week leading up to the MRI scanning session. Session five consisted of a baseline block, three training blocks and a repeat baseline block and was completed on the same day as the MRI scan (just before scanning). Session six consisted of a baseline block only and was completed at 4-week follow up. Every session was computerised and delivered online with participants able to complete it on their phone, tablet or computer. The first session began with a brief narrated psycho-educational slideshow about the role of cognitive biases in depression. Each CBM session took about 8-12 minutes to complete. The stimuli used in CBM were composite images of facial expressions from three exemplar identities taken from the Karolinska Directed Emotional Faces dataset (2) and were identical to our previous CBM and fMRI studies (3, 4). Each face image was morphed from unambiguously happy expression through to unambiguously sad expression in a 15-frame morphed face image continuum (Figure S2).

Baseline blocks

A baseline block consisted of 45 trials (with each face of the 15-frame continuum presented three times). In each trial, images were presented one at a time, in random order, for 150 ms. Stimuli were preceded by a fixation cross which was presented for a random period ranging from 1500 to 2500 ms. After stimuli presentation, and to prevent processing of afterimages, a backward mask of noise was presented for 250 ms. Participants were then prompted to judge the emotion of the facial expression (happy or sad) which remained on the screen until the participant made a response (two alternative forced choice procedure). Baseline blocks determined where in the continuum the participant switched from making primarily ‘happy’ judgements to primarily ‘sad’ judgements (their ‘balance point’). Balance points were calculated as: (number of ‘happy’ judgements / 45 trials) * 15 face stimuli.

Training blocks

A training block consisted of 31 trials, in which unambiguous expressions (images 1-2 and 14-15) were presented once, somewhat ambiguous expressions (images 3-5 and 11-13) were presented twice, and ambiguous faces (images 6-10) were presented three times. After each stimulus was presented, participants were asked to make the same judgement (happy/sad) before been given feedback on screen. In the ‘active’ CBM group, the feedback was tailored such that the two faces closest to the balance point that the participant initially categorised as sad (baseline block) were classed as ‘incorrect’, and it should be categorised as ‘happy’ (see Figure S2). In the ‘sham’ CBM group, training blocks were the same except the feedback given was based directly on the participant’s baseline balance point i.e., the feedback did not attempt to change bias, as it reflected the participant’s response baseline. Our earlier work showed that this procedure was perceptually hard to distinguish from the active condition, and that it had no effect on emotion perception.

Please note that Figure S1 has been removed from the pre-print version on MEDRXIV due to inclusion of experimental stimuli with faces. Please contact the corresponding author to request this figure directly.

**Figure S1. Illustration of active Cognitive Bias Modification.** Top panel shows face stimuli morphed from overtly happy to overtly sad and an example of the balance point measured during a pre-training baseline block. The middle panel shows an example training trial, with timings, where active CBM delivers tailored feedback on the two faces closest to the balance point that the participant judged as ‘sad’ at baseline, whereas sham CBM reinforces initial baseline judgements. The bottom panel shows an example of the balance point measured during a post-training baseline block where categorisations of ambiguous emotional expressions have been shifted so that more are judged as ‘happy’.

**fMRI facial emotional processing task**

Thirteen 30 second blocks of a fixation cross (baseline condition) were interleaved with twelve 30 second blocks of the emotional task (four blocks of sad faces (sad condition), four blocks of happy faces (happy condition) and four blocks of fearful faces (fearful condition). During each emotional block, participants viewed 10 emotional faces (5 female) presented on the screen one-by-one which were all taken from a standard set of pictures of facial affect (5). Each face was presented briefly (500 ms) and participants were asked to report the gender of the face (female/male) via an MRI compatible button box to ensure engagement. Within block interstimulus intervals (ISI) ranged between 2500 and 2900 ms. Stimulus presentation and participant button presses were registered and time-locked to fMRI data using E-Prime. Both accuracy (correct gender discrimination) and reaction times were recorded.

**fMRI motion correction**

Three steps were taken given multiband EPI sequences are particularly sensitive to movement. 1) Framewise Displacement (FD) of the six realignment parameters was calculated (6), 2) scrubbing was performed with Artifact Detection Tool (ART) (7) within the CONN toolbox (version 22.v2407) (8) to identify outlier scans for each participant, and 3) denoising of functional images was applied including high-pass frequency filtering of the BOLD timeseries (9, 10). First, fMRI data quality was checked for movement by calculating Framewise Displacement (FD); the sum of the absolute temporal derivatives of the six rigid-body realignment parameters calculated during preprocessing. For each participant, the mean FD and FD distribution over time was inspected. Participants with mean FD > 0.3mm were deemed to have problematic and unrepairable movement and were excluded from the main analysis. Given that multiband EPI sequences are particularly sensitivity to movement, two additional preprocessing steps were taken to further reduce the impact of movement within the CONN toolbox (version 22.v2407) (8). First, scrubbing was performed with Artifact Detection Tool (ART) based outlier detection (7) to identify outlier scans with FD > 0.9mm or global BOLD signal changes above five standard deviations (intermediate threshold) for each participant (11). Second, a standard denoising pipeline was applied to functional images to regress out potential confounding effects from white matter, CSF, realignment parameters and their first order derivatives, outlier scans from scrubbing step, as well as session effects and their first order derivatives and linear trends (9). High-pass frequency filtering of the BOLD timeseries was also applied above 0.008 Hz (10). Further details of quality control indices for scrubbing and denoising steps are available in the Supplementary results section below.

**Region of Interest Analyses**

For amygdalae, binary masks were anatomically defined using the probabilistic Harvard Oxford Subcortical structural atlas (voxels >50% probability). Left and right amygdalae were merged into a single ROI mask. For the mPFC, peak coordinates (MNI 8, 56, 18) were taken from a recent meta-analysis showing mPFC activation by psychotherapy in depression (Nord et al., 2021). A 10mm sphere around the peak coordinates was used.

**Functional connectivity (PPI)**

The seed region was defined based on peak activation in the mPFC (MNI coordinates: 14, 62, 14) at a threshold of p<0.001 uncorrected, with a 10mm radius sphere. The deconvolved BOLD signal from all voxels within this seed region sphere was extracted for each participant (physiological regressor) in response to happy, fearful and sad emotions (psychological regressors). Three interaction terms (PPI regressors) were computed as the element-wise multiplication of the mean-centred physiological and psychological variables (happy PPI, fearful PPI and sad PPI). These three regressors (one physiological, three psychological, three PPI interaction terms) were entered into a first-level GLM for each participant along with the original motion parameters and subject-specific outlier scans. A second-level random effects analysis using a full factorial model was then constructed to test the effect of group (active/sham CBM) on PPI regressors. Additionally, full factorial models were also constructed with individual change in balance point scores (from pre-training to scanning session, and to 4-week follow-up) added as covariates and set to interact with CBM group. This was to investigate any group x change in balance point interactions on mPFC connectivity.

**Results**

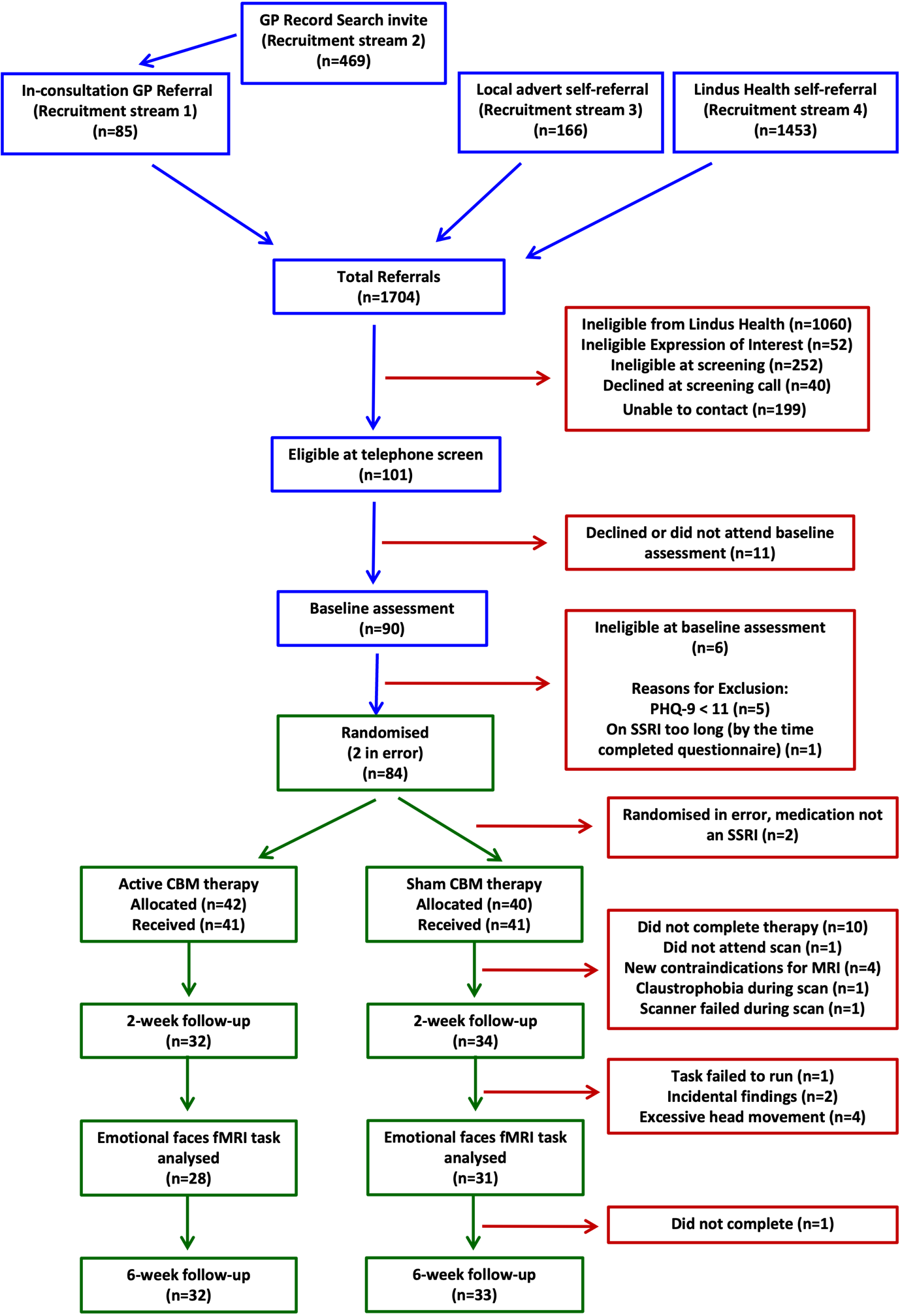

**Figure S2. CONSORT diagram of recruitment.**

**FMRI data quality control – movement**

Denoising:

Functional data were denoised using a standard denoising pipeline including the regression of potential confounding effects characterized by white matter timeseries (5 CompCor noise components), CSF timeseries (5 CompCor noise components), motion parameters and their first order derivatives (12 factors), outlier scans (below 76 factors), session effects and their first order derivatives (2 factors), and linear trends (2 factors) within each functional run, followed by high-pass frequency filtering of the BOLD timeseries above 0.008 Hz. CompCor noise components within white matter and CSF were estimated by computing the average BOLD signal as well as the largest principal components orthogonal to the BOLD average, motion parameters, and outlier scans within each subject's eroded segmentation masks. From the number of noise terms included in this denoising strategy, the effective degrees of freedom of the BOLD signal after denoising were estimated to range from 23.4 to 501.7 (average 459.8) across all subjects.

Quality control indices for motion correction steps:

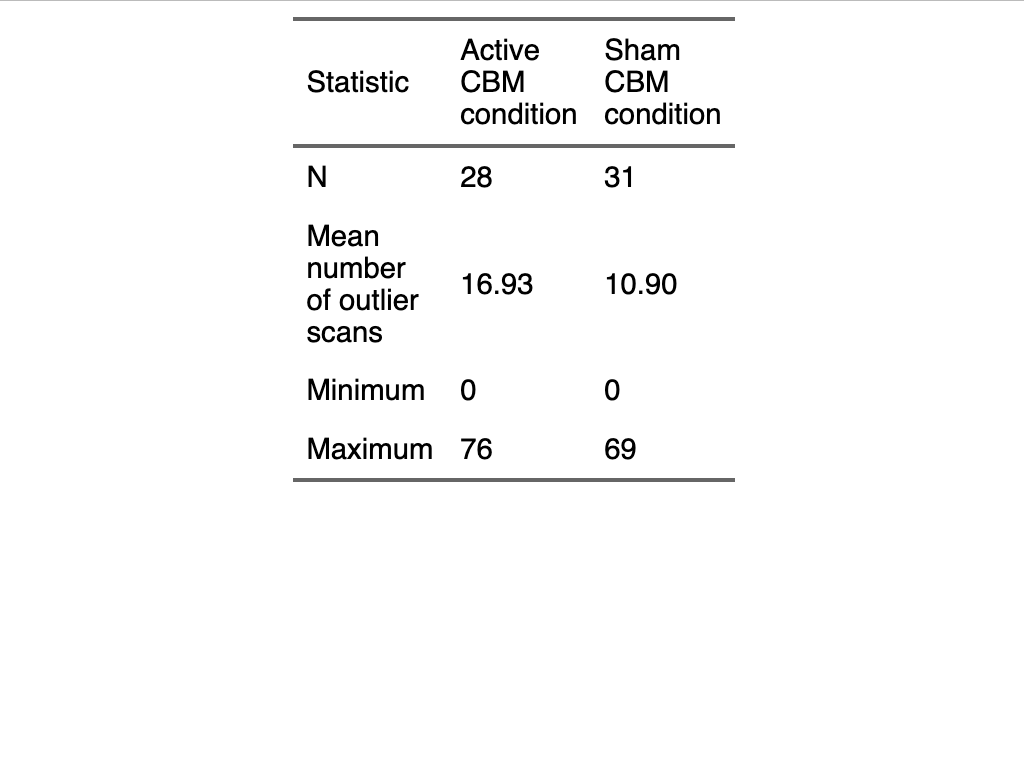

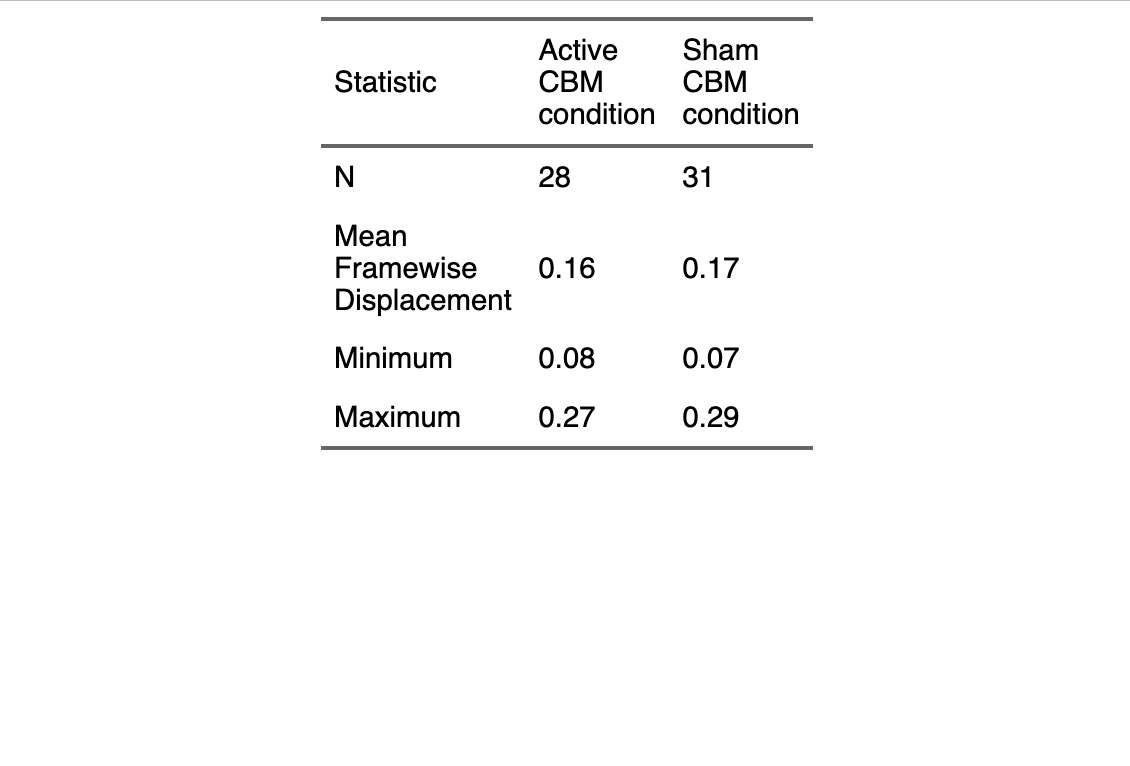

**Table S1.** Mean framewise displacement (from realignment preprocessing step) and mean number of outlier scans (from scrubbing preprocessing step) between active and sham CBM groups for n=59 participants included in final fMRI data analysis. There were no significant differences between groups. Five participants with mean framewise displacement above 0.3mm were excluded from the sample.

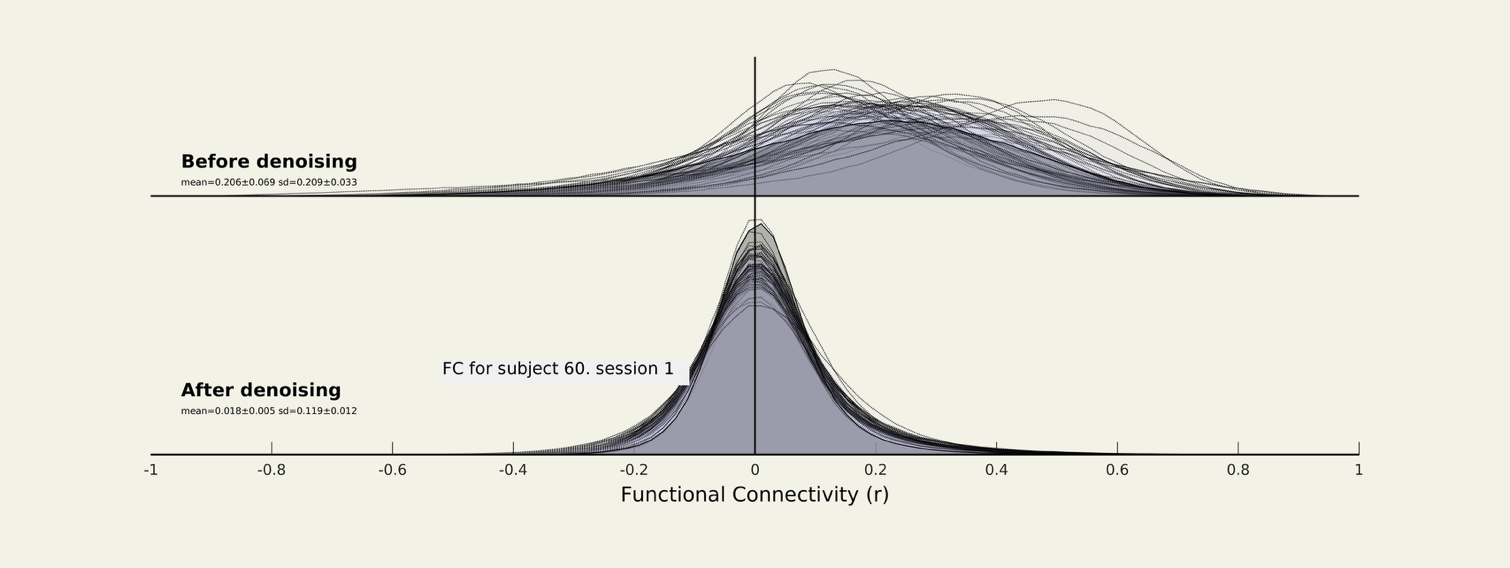

**Figure S3. Quality Control plots from denoising step in CONN toolbox.** Plots are histograms of functional correlation values between pairs of voxels before and after denoising, where each histogram represents one participant. The denoising step improves the distribution of these functional connectivity values.

**Clinical outcome measures**

There was strong evidence that all participants showed improvements in depressive symptoms (mean Difference = -5.40), anxiety (mean difference = -4.66), panic (mean difference = -1), anhedonia (mean difference = 5.04) and quality of life (mean difference = 6.42) from baseline to 4-week follow up (Figures S4-S8 below). There was weak evidence that the active CBM group had a larger improvement in panic symptoms from baseline to scanning session (beta = -0.99 [95% CI = -1.86 - -0.14], *p*=.023) and 4-week follow up (beta = -0.85 [95% CI = -1.75 – 0.04], *p*= .063), but not for any other clinical measures.

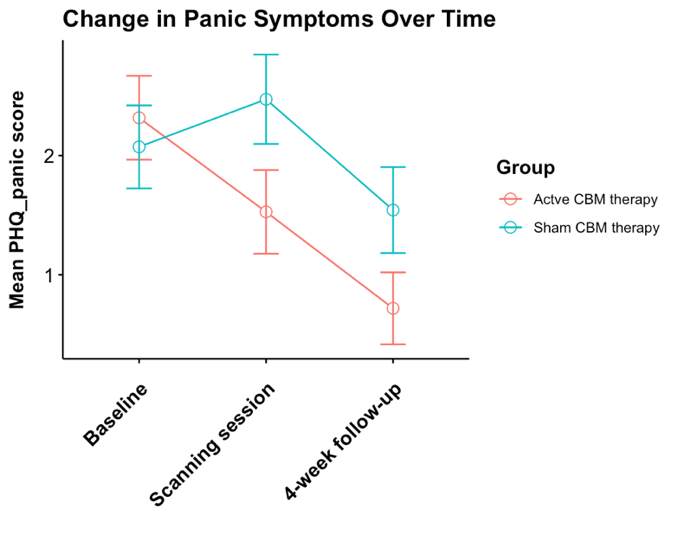
**Figures S4-S8:**
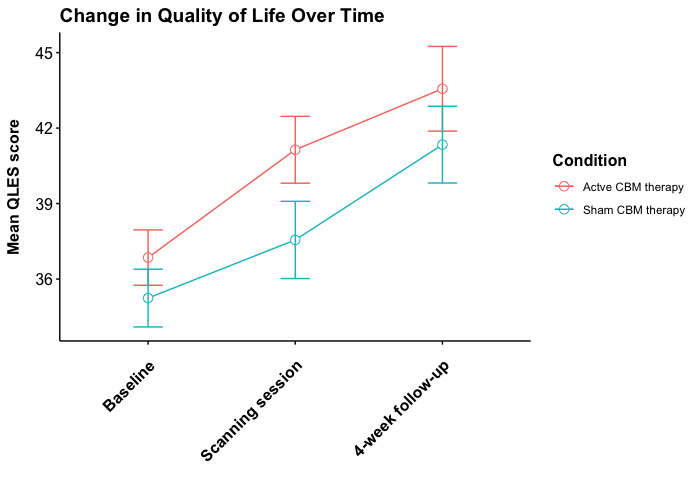

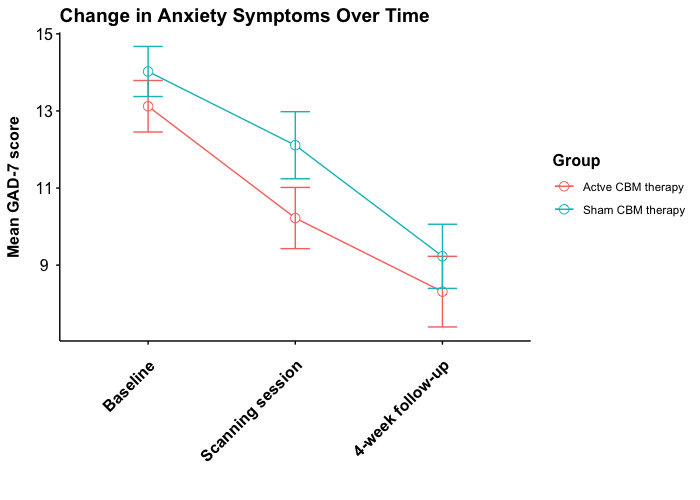

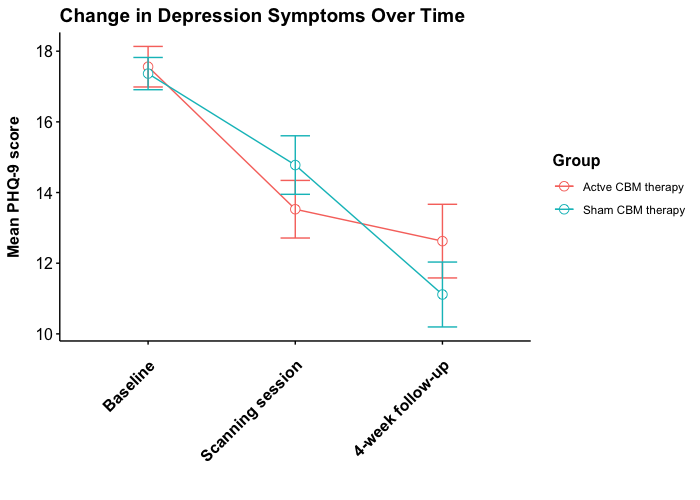

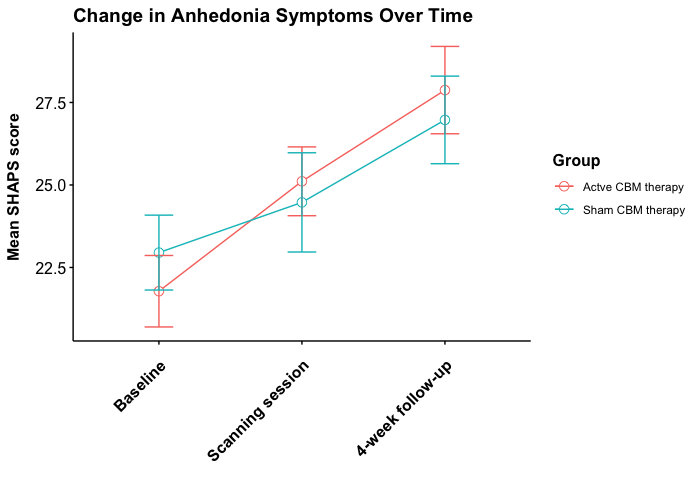

| Brain region | Cluster-level statistics | | Peak-level statistics | | MNI coordinates | | |
| --- | --- | --- | --- | --- | --- | --- | --- |
|  | FWE-corrected p-value | K | FWE-corrected p-value | F value | X | Y | Z |
| Left Superior Temporal Gyrus | <.001 | 10466 | <.001 | 404.95 | 40 | -44 | -24 |
| Left Middle Temporal Gyrus |  |  | <.001 | 401.64 | 26 | -50 | -22 |
| Left Inferior Temporal Gyrus |  |  | <.001 | 276.38 | -38 | -48 | -22 |
| Right Supplementary Motor Area | <.001 | 1719 | <.001 | 335.5 | -2 | 4 | 54 |
| Left Cerebellum | <.001 | 750 | <.001 | 254.19 | 18 | -62 | -48 |
| Left Cerebellum |  |  | <.001 | 69.69 | 8 | -74 | -42 |
| Right Precuneus | <.001 | 5633 | <.001 | 244.61 | -34 | -10 | 66 |
| Right Precuneus |  |  | <.001 | 244.14 | -46 | -26 | 58 |
| Right Precuneus |  |  | <.001 | 237.5 | -38 | -26 | 52 |
| Right Precentral Gyrus | <.001 | 18179 | <.001 | 224.95 | 40 | -18 | 48 |
| Left Angular Gyrus |  |  | <.001 | 217.69 | 20 | -58 | 20 |
| Left Angular Gyrus |  |  | <.001 | 203.07 | -28 | -42 | -8 |
| Left Occipital Cortex (Lingual Gyrus) | <.001 | 158 | <.001 | 198.97 | -30 | -92 | -6 |
| Left Insula / Putamen | <.001 | 156 | <.001 | 128.25 | -14 | -18 | 8 |
| Left Putamen / Globus Pallidus |  |  | 0.017 | 29.8 | -18 | -16 | 18 |
| Right Occipital Cortex (Calcarine) | <.001 | 200 | <.001 | 126.97 | 26 | -96 | -6 |
| Left Superior Temporal Gyrus | <.001 | 2163 | <.001 | 123.1 | -60 | -28 | 6 |
| Left Middle Temporal Gyrus |  |  | <.001 | 117.16 | -42 | -18 | -4 |
| Left Superior Temporal Gyrus |  |  | <.001 | 94.35 | -58 | -8 | -6 |
| Left Cerebellum | <.001 | 552 | <.001 | 119.46 | -12 | -50 | -48 |
| Right Cerebellum |  |  | <.001 | 114.48 | 12 | -48 | -46 |
| Right Cerebellum |  |  | <.001 | 65.94 | 10 | -50 | -54 |
| Right Cerebellum | <.001 | 210 | <.001 | 109.35 | 30 | -40 | -10 |
| Left Precentral Gyrus | <.001 | 1226 | <.001 | 108.14 | -22 | 14 | 50 |
| Left Superior Frontal Gyrus |  |  | <.001 | 101.93 | -24 | 30 | 40 |
| Right Superior Frontal Gyrus | <.001 | 712 | <.001 | 102.66 | 26 | 26 | 44 |
| Right Superior Frontal Gyrus |  |  | <.001 | 61.8 | 30 | 32 | 52 |
| Right Precentral Gyrus |  |  | <.001 | 48.61 | 24 | 14 | 54 |
| Left Cerebellum | <.001 | 235 | <.001 | 93.5 | -32 | -66 | -52 |
| Left Supplementary Motor Area | <.001 | 2819 | <.001 | 92.84 | -10 | 46 | -4 |
| Right Supplementary Motor Area |  |  | <.001 | 89.7 | 10 | 44 | -6 |
| Right Middle Frontal Gyrus |  |  | <.001 | 79.09 | 6 | 34 | 12 |
| Right Postcentral Gyrus | <.001 | 2444 | <.001 | 85.28 | 38 | -2 | 60 |
| Right Middle Frontal Gyrus |  |  | <.001 | 84.85 | 34 | 18 | 6 |
| Right Precentral Gyrus |  |  | <.001 | 83.86 | 42 | 0 | 42 |
| Right Angular Gyrus | <.001 | 603 | <.001 | 74.71 | 34 | -56 | 42 |
| Right Supramarginal Gyrus |  |  | <.001 | 45.05 | 46 | -36 | 46 |
| Left Superior Frontal Gyrus | <.001 | 114 | <.001 | 59 | -26 | 36 | -12 |
| Left Middle Temporal Gyrus | <.001 | 125 | <.001 | 55.73 | -56 | -60 | -10 |
| Right Fusiform Gyrus | <.001 | 22 | <.001 | 55.44 | 32 | -8 | -34 |
| Right Cerebellum | <.001 | 126 | <.001 | 49.37 | 46 | -56 | -42 |
| Right Cerebellum |  |  | 0.005 | 32.77 | 40 | -70 | -40 |
| Right Hippocampus | <.001 | 32 | <.001 | 48.96 | 18 | -6 | -14 |
| Right Superior Frontal Gyrus | <.001 | 27 | <.001 | 48.78 | 22 | 60 | 0 |
| Left Occipital Cortex | <.001 | 23 | <.001 | 44.19 | -38 | -72 | 6 |
| Right Cerebellum | <.001 | 24 | <.001 | 43.72 | 10 | -30 | -34 |
| Right Cerebellum | <.001 | 94 | <.001 | 43.35 | 30 | -18 | -20 |
| Right Cerebellum |  |  | 0.001 | 37.46 | 20 | -16 | -24 |
| Right Cerebellum | <.001 | 30 | <.001 | 41.02 | 16 | -82 | -40 |
| Right Middle Temporal Gyrus | <.001 | 33 | <.001 | 41.01 | 22 | 42 | -14 |
| Right Middle Occipital Cortex | 0.001 | 15 | <.001 | 40.59 | 42 | -66 | 2 |
| Left Putamen | 0.003 | 7 | 0.004 | 33.72 | -34 | -18 | 18 |
| Left Amygdala | 0.001 | 14 | 0.005 | 33.09 | -16 | -10 | -14 |
| Left Cerebellum | 0.002 | 9 | 0.005 | 32.94 | -46 | -60 | -40 |
| Right Hippocampus | 0.008 | 4 | 0.009 | 31.43 | 12 | -14 | 8 |
| Right Superior Temporal Gyrus | 0.002 | 9 | 0.011 | 31.08 | 56 | -38 | -14 |
| Right Middle Temporal Gyrus | 0.004 | 6 | 0.011 | 31.01 | 58 | -40 | 24 |
| Left Insula | 0.016 | 2 | 0.016 | 30.01 | -28 | -20 | 4 |
| Right Middle Frontal Gyrus | 0.008 | 4 | 0.016 | 29.95 | 36 | 42 | -12 |
| Right Superior Temporal Gyrus | 0.006 | 5 | 0.019 | 29.57 | 66 | -38 | -6 |
| Left Insula | 0.016 | 2 | 0.021 | 29.36 | -34 | 12 | -12 |
| Left Cerebellum | 0.024 | 1 | 0.022 | 29.21 | -34 | -10 | -30 |
| Right Insula | 0.024 | 1 | 0.035 | 27.95 | 24 | -4 | 12 |

**Table S2.** Pattern of neural activation evoked during the task (average effect of viewing happy, fearful and sad faces compared to implicit baseline) across groups. Results were thresholded at p=.05 FWE-corrected and both cluster-level and peak-level statistics are presented along with MNI coordinates. K = number of voxels in cluster.

**fMRI imaging sensitivity analyses**

Type of SRRI medication. To understand the effect of different types of SSRI medication, we conducted a sensitivity analysis on our primary analyses restricted only to patients taking Sertraline which was the most commonly prescribed SSRI antidepressant (n=36). Although these unadjusted analyses were very underpowered, the peak activations in right IOG (main effect of CBM therapy group) and mPFC (group x happy-fear contrast) were qualitatively similar to the findings reported in the full sample (n=59) although the point estimates were slightly reduced (mPFC: T=2.92, p=.164, peak = 12, 56, 16; Right IOG: T=3.37, p=1 FWE corrected, peak = 24, 092, 2).

Medication Duration. To assess the impact of medication duration on the primary fMRI findings (group x happy-fearful faces interaction in mPFC), differences in mPFC activity between groups for this contrast were visually inspected for participants with ‘longer’ (2-6 months) and ‘shorter’ (0-2 months) medication duration at baseline assessment (see Figure below). Medication duration categories were based on a median split of the data where there was n= 30 in the longer group, and n= 26 in the shorter group (n=3 did not have this data available). mPFC activity appears most different between CBM groups in participants with a longer medication duration at baseline, than shorter medication duration, although no inferential statistics were performed due to the exploratory, and underpowered, nature of this analysis. There was therefore no evidence that a sensitive window exists to intervene with CBM therapy in the first few weeks of SSRI treatment.

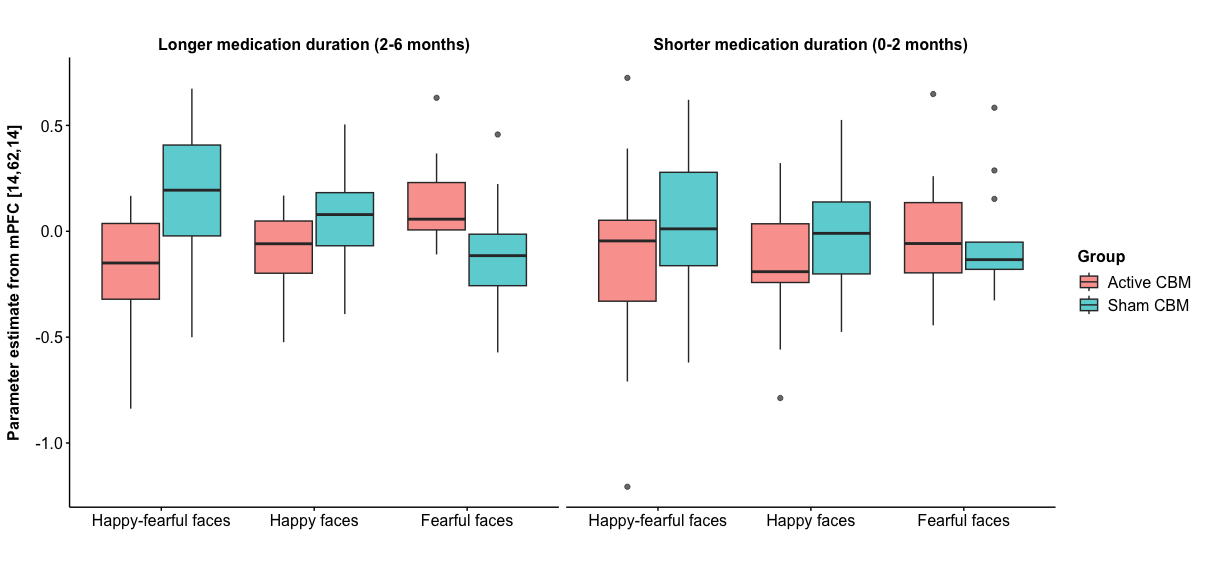

**Figure S9. Effect of medication duration on group x happy-fearful contrast interaction.** Box plots are presented showing parameter estimates taken from peak mPFC voxel for happy-fearful faces contrast as well as in response to happy and fearful faces separately, split by medication duration (longer/shorter).

Impact of Age. To assess the impact of age on the primary fMRI findings (group x happy-fearful faces interaction in mPFC), differences in mPFC activity between groups for this contrast were visually inspected for ‘older’ (31-50) and ‘younger’ (19-30) participants (see Figure below). Age categories were based on a median split of the data where there was n= 30 in the older group, and n= 29 in the younger group. mPFC activity appears most different between CBM groups in older participants, than younger participants, although no inferential statistics were performed due to the exploratory, and underpowered, nature of this analysis. There was therefore no evidence that CBM therapy may be more effective in younger adults.

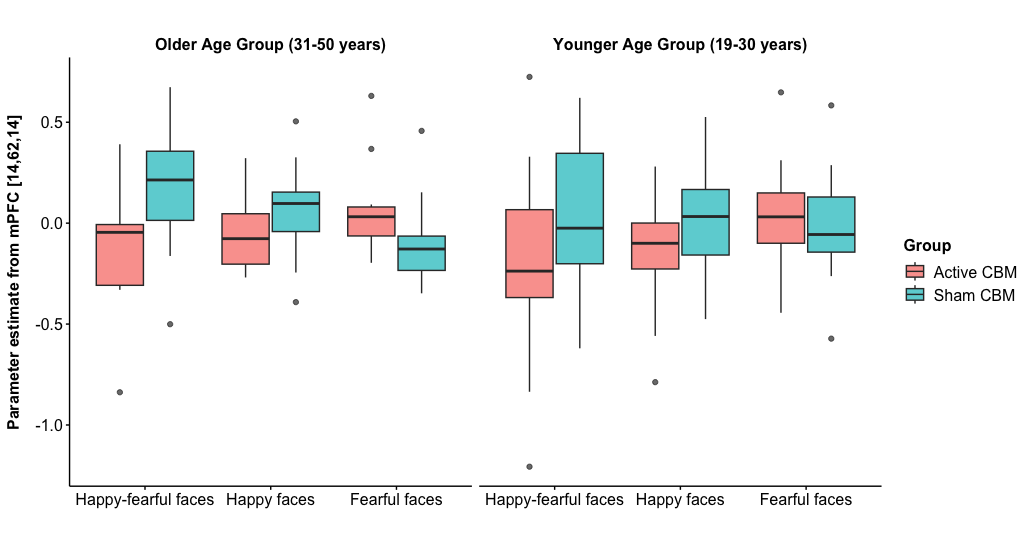

**Figure S10. Effect of age on group x happy-fearful contrast interaction.** Box plots are presented showing parameter estimates taken from peak mPFC voxel for happy-fearful faces contrast as well as in response to happy and fearful faces separately, split by age group (older/younger).

**Further references**

1.	Kroenke K, Spitzer R, Williams J. The PHQ-9: validity of a brief depression severity measure. J. Gen. Intern. Med. 2001;16(9):606-13.

2.	Lundqvist D, Flykt A, Öhman A. Karolinska directed emotional faces. Cognition and Emotion. 1998.

3.	Penton-Voak IS, Bate H, Lewis G, Munafò MR. Effects of emotion perception training on mood in undergraduate students: randomised controlled trial. Br J Psychiatry. 2012;201(1):71-2. doi:10.1192/bjp.bp.111.107086

4.	Penton-Voak IS, Adams S, Button KS, Fluharty M, Dalili M, Browning M, et al. Emotional recognition training modifies neural response to emotional faces but does not improve mood in healthy volunteers with high levels of depressive symptoms. Psychol Med. 2021;51(7):1211-9. doi:10.1017/s0033291719004124

5.	Tottenham N, Tanaka JW, Leon AC, McCarry T, Nurse M, Hare TA, et al. The NimStim set of facial expressions: Judgments from untrained research participants. Psychiatry Research. 2009;168(3):242-9. doi:10.1016/j.psychres.2008.05.006

6.	Power JD, Barnes KA, Snyder AZ, Schlaggar BL, Petersen SE. Spurious but systematic correlations in functional connectivity MRI networks arise from subject motion. NeuroImage. 2012;59(3):2142-54. doi:10.1016/j.neuroimage.2011.10.018

7.	Morfini F, Whitfield-Gabrieli S, Nieto-Castañón A. Functional connectivity MRI quality control procedures in CONN. Front Neurosci. 2023;17:1092125. doi:10.3389/fnins.2023.1092125

8.	Whitfield-Gabrieli S, Nieto-Castanon A. Conn: a functional connectivity toolbox for correlated and anticorrelated brain networks. Brain Connect. 2012;2(3):125-41. doi:10.1089/brain.2012.0073

9.	Power JD, Mitra A, Laumann TO, Snyder AZ, Schlaggar BL, Petersen SE. Methods to detect, characterize, and remove motion artifact in resting state fMRI. Neuroimage. 2014;84:320-41. doi:10.1016/j.neuroimage.2013.08.048

10.	Hallquist MN, Hwang K, Luna B. The nuisance of nuisance regression: spectral misspecification in a common approach to resting-state fMRI preprocessing reintroduces noise and obscures functional connectivity. Neuroimage. 2013;82:208-25. doi:10.1016/j.neuroimage.2013.05.116

11.	Ashburner J, editor. Preparing fMRI Data for Statistical Analysis2009.
